# Genetic associations of protein-coding variants in human disease

**DOI:** 10.1101/2021.10.14.21265023

**Authors:** Benjamin B. Sun, Mitja I. Kurki, Christopher N. Foley, Asma Mechakra, Chia-Yen Chen, Eric Marshall, Jemma B. Wilk, Biogen Biobank Team, Mohamed Chahine, Philippe Chevalier, Georges Christé, FinnGen, Aarno Palotie, Mark J. Daly, Heiko Runz

## Abstract

Genome-wide association studies (GWAS) have identified thousands of genetic variants linked to the risk of human disease. However, GWAS have thus far remained largely underpowered to identify associations in the rare and low frequency allelic spectrum and have lacked the resolution to trace causal mechanisms to underlying genes. Here, we combined whole exome sequencing in 392,814 UK Biobank participants with imputed genotypes from 260,405 FinnGen participants (653,219 total individuals) to conduct association meta-analyses for 744 disease endpoints across the protein-coding allelic frequency spectrum, bridging the gap between common and rare variant studies. We identified 975 associations, with more than one-third of our findings not reported previously. We demonstrate population-level relevance for mutations previously ascribed to causing single-gene disorders, map GWAS associations to likely causal genes, explain disease mechanisms, and systematically relate disease associations to levels of 117 biomarkers and clinical-stage drug targets. Combining sequencing and genotyping in two population biobanks allowed us to benefit from increased power to detect and explain disease associations, validate findings through replication and propose medical actionability for rare genetic variants. Our study provides a compendium of protein-coding variant associations for future insights into disease biology and drug discovery.

## Introduction

Inherited protein-coding and non-coding DNA variations play a role in the risk, onset, and progression of human disease. Traditionally, geneticists have dichotomized diseases as either caused by coding mutations in single genes that tend to be rare, highly penetrant, and often compromise survival and reproduction (often termed “Mendelian” diseases), or alternatively as common diseases that show a complex pattern of inheritance influenced by the joint contributions of hundreds of low-impact, typically non-coding genetic variants (often termed “complex” diseases). For both rare and common conditions, large human cohorts systematically characterized for a respective trait of interest have enabled the identification of thousands of disease-relevant variants through either sequencing-based approaches or genome-wide association studies (GWAS). Nevertheless, the exact causal alleles and mechanisms that underlie associations of genetic variants to disease have thus far remained largely elusive^1^.

In recent years, population biobanks have been added to the toolkit for disease gene discovery. Biobanks provide the opportunity to simultaneously investigate multiple traits and diseases at once and uncover relationships between previously unconnected phenotypes. For instance, the UK Biobank (UKB) is a resource that captures detailed phenotype information matched to genetic data for over 500,000 individuals and, since its inception, has facilitated biomedical discoveries at an unprecedented scale^2^. We and others have recently reported on the ongoing efforts to sequence the exomes of all UKB participants and link genetic findings to a broad range of phenotypes^3-5^. We also established FinnGen (FG), an academic-industry collaboration to identify genotype-phenotype correlations in the Finnish founder population with the aim to better understand how the genome affects health (https://www.finngen.fi). Finland is a well-established genetic isolate and a unique gene pool distinguishes Finns from other Europeans^6^. The distinct Finnish haplotype structure is characterized by large blocks of co-inherited DNA in linkage disequilibrium and an enrichment for alleles that are rare in other populations, but can still be confidently imputed from genotyping data even in the rare and ultra-rare allele frequency spectrum^7-9^. Through combining imputed genotypes with detailed phenotypes ascertained through national registries, FG holds the promise to provide particular insights into the yet little examined allele frequency spectrum between 0.1 and 2% where both sequencing studies and GWAS have thus far remained largely underpowered to identify associations to disease. This spectrum includes many coding variants with moderate to large effect sizes that can help identify causal genes in GWAS loci, provide mechanistic insights into disease pathologies, and potentially bridge rare and common diseases.

Here, we have leveraged the combined power of UKB and FG to investigate how rare and low-frequency variants in protein-coding regions of the genome contribute to the risk for human traits and diseases. Using data from a total of 653,219 individuals, we tested how ∼48,000 coding variants identified in both biobanks through either whole-exome sequencing (WES) or genotype imputation associate with 744 distinct disease endpoints. Disease associations were compared against information from rare disease, biomarker and drug target resources and complemented by deep dives into distinct disease mechanisms of individual genes and coding variants. Our results showcase the benefits of combining large population cohorts to discover and replicate novel associations, explain disease mechanisms across a range of common and rare diseases, and shed light on a substantial gap in the allelic spectrum that neither genotyping nor sequencing studies have previously been able to address.

## Results

An overview of the study design and basic demographics are provided in **Extended Data Figure 1 and Supplementary Table 1**. In brief, we systematically harmonised disease phenotypes across UKB and FG using Phecode and ICD10 mappings and retained 744 specific disease endpoints grouped into 580 disease clusters that span a broad range of diseases (**Methods, Supplementary Table 2**). Disease case counts relative to cohort size showed good correlations both, overall between UKB and FG (Spearman’s =0.65, *p*<5.3×10^−90^) and across distinct disease groups (**Extended Data Figure 2**).

### Coding-wide association analyses in 653,219 individuals across 744 disease endpoints identify 975 genetic signals

We performed coding-wide association studies (CWAS) across 744 disease endpoints over a mean of 48,189 (range: 25,309-89,993, **Methods, Supplementary Table 2**) post-QC coding variants across the allele frequency spectrum derived from whole-exome sequencing (WES) of 392,814 European ancestry individuals in UKB and meta-analysed these data with summary results from up to 260,405 individuals in FG (**Methods, Supplementary Table 2**).

We identified 975 associations (534 variants in 301 distinct regions across 148 disease clusters; 620 distinct region-disease cluster associations) meeting genome-wide significance (*p*<5×10^−8^), and 717 associations (378 variants in 231 distinct regions across 121 disease clusters; 445 distinct region-disease cluster associations) at a conservative (Bonferroni) multiple testing threshold of *p*<2×10^−9^ (correcting for the number of approximate independent tests) (**Methods, Figure 1a, Supplementary Figure 1 (interactive), Supplementary Table 3**). The distributions of coding variant annotation categories were largely similar for variants with at least one significant association (*p*<5×10^−8^) relative to all variants tested, with missense variants showing a higher fraction of significant variants than in-frame indel or predicted loss-of-function (pLoF) variants (**Extended Data Figure 3**). Inflation was well controlled with a mean genomic inflation factor of 1.04 (5-95 percentiles: 1.00-1.09, **Extended Data Figure 4a**). Effect sizes were generally well aligned between UKB and FG (Spearman’s =0.90, *p*<10^−300^, **Extended Data Figure 4b**). MAFs of lead variants correlated well overall between UKB and FG (Spearman’s =0.97, *p*<10^−300^, **Figure 1b**), especially for variants with MAF>1%, yet as expected^8^ from genetic differences between Finns and non-Finnish Europeans (NFEs) was reduced for variants with MAF<1% (Spearman’s =0.32, *p*=0.023).

**Figure 1.**
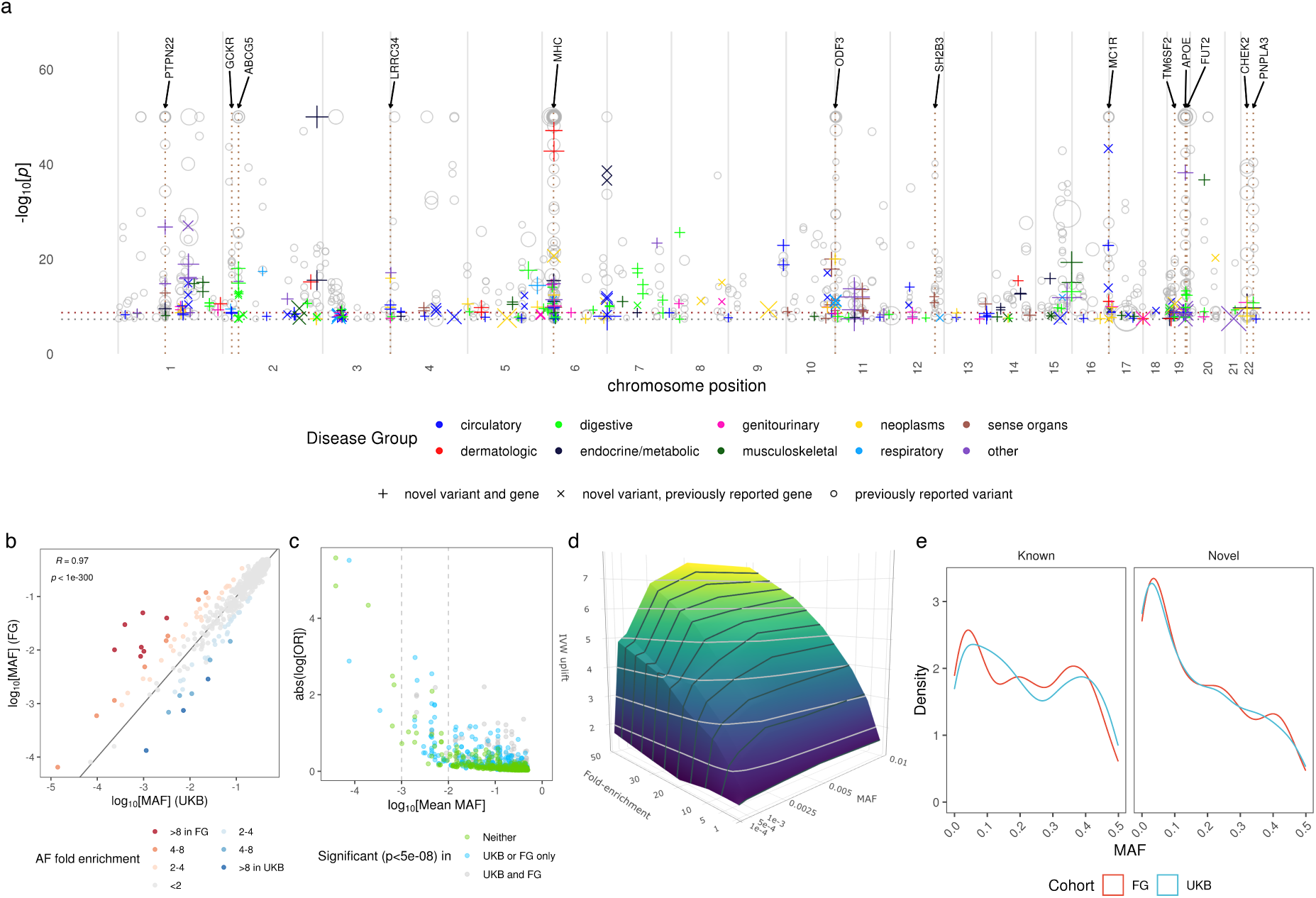
Coding genetic associations with disease. Manhattan plot for novel associations and allelic enrichment surface plots are provided as Interactive Supplementary Figures 1 and 2. **(a) Summary of sentinel variant associations.** Size of the point is proportional to effect size. -log_10_(*p*) capped at -log_10_(10^−50^). Labels highlight pleiotropic associations (≥ 5 trait clusters). Colours indicate disease groups. Shape indicate novel/known (grey circles) associations. Dotted horizontal lines: - log_10_(2×10^−9^) [brown] and -log_10_(5×10^−8^) [grey]. **(b). Comparison of sentinel variant MAF between UKB and FG. (c) Effect size against MAF of sentinel variants**. Dashed lines indicate MAF of 0.1% (left) and 1% (right). **(d) Surface plot of effects of cohort specific allele enrichment on inverse variant weighted meta-analysis z-scores (IVW uplift) across MAFs** (up to MAF 1%). Uplift is defined as the ratio of meta-analysed IVW Z-score to the Z-score of an individual study (details in **Supplementary Information**). **(e) Density plot of MAF for sentinel variants for known vs novel associations**.

Across all diseases, we found generally larger effect sizes for low frequency and rare variants (**Figure 1c**). 387 of the 975 (39.7% at *p*<5×10^−8^; 270/717 (37.7%) at *p*<2×10^−9^) associations would not have been detected if analysed in UKB (61.5% at *p*>5×10^−8^; 60.1% at *p*>2×10^−9^) or FG (59.6% at *p*>5×10^−8^; 58.6% at *p*>2×10^−9^) alone. Association testing within UKB and FG individually would have yielded 318 and 479 associations respectively at *p*<5×10^−8^ **(Supplementary Tables 4 and 5)**. Thus our combined approach utilizing both biobanks increased the number of significant findings by approximately 3- and 2-fold, respectively. Of the 318 and 479 significant sentinel variants in UKB and FG, 252 (72.6%) and 258 (53.9%) replicated at *p*<0.05 in FG and UKB respectively **(Supplementary Tables 4 and 5)**, highlighting further the strength of our approach to yield results that are more robust through replication than would be findings derived from just a single biobank.

Our study benefits from population enrichment of rare alleles in Finns versus NFEs (and vice versa) that increases the power for association discovery. Using a combination of theoretical analyses and empirical simulations, we show that by leveraging population-enriched variants we could increase inverse-variance weighted meta-analysis Z-scores and hence our ability to detect underlying associations. The gain in power from enriched alleles was present across a range of rare MAFs (0.01-1%), with the strongest power gain in the rare and ultra-rare minor allele frequency (MAF) range of 0.01% to 0.25% (**Figure 1d, Supplementary Information, Extended Data Figure 5, Supplementary Figure 2 (interactive))**. Of the sentinel variants, we found 73 (33 in UKB, 40 in FG) to be enriched by >2-fold and 23 (8 in UKB, 15 in FG) by >4-fold relative to the respective other biobank (**Figure 1b, Supplementary Table 6**). The majority of highly population enriched variants are rare (MAF<1%) or low frequency (MAF 1-5%), whereby 20 of 23 variants with >4-fold population enrichment (13 in FG and 7 in UKB) had MAF <1% (**Table 1, Supplementary Table 6**).

**Table 1.** Genes with sentinel variants enriched >4 fold in either UKB or FG. All enrichment *p*<5×10^−5^.

| Gene | rsID (protein change) | CHR | A0/A1 | A1 freq UKB/FG% | log2 FE (FG/UKB) | OMIM gene phenotype relationships | CWAS gene phenotype relationships |
| --- | --- | --- | --- | --- | --- | --- | --- |
| *CHEK2* | rs17879961 (I200T)<br>rs555607708<sup>1</sup> (T410fs) | 22 | A/G<br>AG/- | 0.04/2.99%<br>0.24/0.64% | 6.25<br>1.42 | Cancer (breast, prostate, colorectal, osteosarcoma); Li-Fraumeni syndrome | rs17879961:G – benign meningeal neoplasm<br>rs555607708:del (2.8x FG enriched) – Cancer (breast, thyroid, colorectal (benign)); uterine leiomyoma; ovarian cysts; PCOS |
| *DBH* | rs77273740 (R79W) | 9 | C/T | 0.10/4.95% | 5.69 | Orthostatic hypotension | Hypertension (IA) |
| *PITX2* | rs143452464 (P41S) | 4 | G/A | 0.02/1.01% | 5.42 | Anterior segment dysgenesis; Axenfeld-Rieger syndrome, ring dermoid of cornea | Arrhythmia and AF |
| *SLC24A5* | rs1426654 (T111A) | 15 | A/G | 0.09/1.13% | 3.67 | Skin/hair/eye pigmentation (dark); oculocutaneous albinism | Non-epithelial cancer of skin (other) (IA) |
| *CFHR5* | rs565457964 (E163fs) | 1 | C/CAA | 0.32/3.96% | 3.66 | Nephropathy due to CFHR5 deficiency | Degeneration of macula and posterior pole of retina (IA) |
| *ANKH* | rs146886108 (R187Q) | 5 | C/T | 0.72/0.07% | -3.28 | Chondrocalcinosis; craniometaphyseal dysplasia | Type 2 diabetes mellitus (IA) |
| *ALDH16A1* | rs150414818 (P527R) | 19 | C/G | 0.10/0.95% | 3.23 | - | Gout |
| *LRRK1* | rs41531245 (T967M) | 15 | C/T | 0.09/0.76% | 3.15 | - | Contracture of palmar fascia; fasciitis; umbilical hernia |
| *CFI* | rs141853578 (G119R) | 4 | C/T | 0.11/0.01% | -3.10 | Atypical haemolytic uremic syndrome; age-related macular degeneration; CFI deficiency | Retinal disorders (other) |
| *FLG* | rs61816761 (R501*)<br>rs138381300<sup>1</sup> (S761fs) | 1 | G/A<br>CACTG/- | 2.45/0.29%<br>2.45/1.35% | -3.10<br>-0.85 | Atopic dermatitis; ichthyosis vulgaris | rs61816761:A – dermatitis (other)<br>rs138381300:del (1.8x UKB enriched) – asthma; non-epithelial cancer of skin (other) |
| *SOS2* | rs72681869 (P191R) | 14 | G/C | 1.09/0.15% | -2.84 | Noonan syndrome | Hypertension (IA) |
| *XPA* | rs144725456 (H244R) | 9 | T/C | 0.01/0.06% | 2.61 | Xeroderma pigmentosum | Non-epithelial cancer of skin (other) |
| *CDC25A* | rs146179438 (Q24H) | 3 | C/A | 1.52/8.72% | 2.52 | - | Kidney and urinary stones (IA) |
| *F10* | rs61753266 (E142K) | 13 | G/A | 0.33/1.83% | 2.46 | Factor X deficiency | PE and pulmonary heart disease (inverse association) |
| *TNXB* | rs61745355 (G2848R)<br>rs10947230<sup>1</sup> (R2704H)<br>rs1150752<sup>1</sup> (T302A) | 6 | C/T<br>C/T<br>T/C | 2.22/11.86%<br>5.96/14.75%<br>13.29/9.17% | 2.42<br>1.31<br>-0.54 | Ehlers-Danlos syndrome; vesicoureteral reflux | rs61745355:T – lymphoma<br>rs10947230:T – lichen planus<br>rs1150752:C – chronic hepatitis; other inflammatory liver diseases; atherosclerosis |
| *SLC39A8* | rs13107325 (A391T) | 4 | C/T | 7.40/1.46% | -2.35 | Congenital disorder of glycosylation | Shoulder lesions |
| *CLPTM1* | rs150484293 (L140F) | 19 | C/T | 0.35/0.07% | -2.33 | - | Dementia |
| *ELL2* | rs141299831 (S18L) | 5 | G/A | 0.02/0.12% | 2.29 | - | Benign neoplasm of other and ill-defined parts of digestive system |
| *CASP7* | rs141266925 (F214L) | 10 | T/C | 0.31/1.5% | 2.29 | - | Cataracts |
| *BRCA1* | rs80357906 (Q1777fs) | 17 | T/TG | 0.001/0.01% | 2.21 | Cancer (breast, ovarian, pancreatic); Fanconi anaemia | Breast cancer |
| *SCN5A* | rs45620037 (T220I) | 3 | G/A | 0.11/0.49% | 2.20 | Sudden infant death syndrome; dilated cardiomyopathy; arrhythmia<sup>2</sup> | Arrhythmia and AF |
| *CACNA1D* | rs1250342280 (F1943del) | 3 | CCTT/C | 0.60/0.14% | -2.09 | Primary aldosteronism, seizures, and neurologic abnormalities; sinoatrial node dysfunction and deafness | Hypertension |
| *WNT10A* | rs121908120 (F228I) | 2 | T/A | 2.72/0.65% | -2.06 | Odontoonychodermal dysplasia; Schopf-Schulz-Passarge syndrome; selective tooth agenesis | Follicular cysts of skin and subcutaneous tissue (IA) |

<sup>1</sup>Other sentinel variants in the gene with <4 FE
<sup>2</sup>Sudden infant death syndrome; atrial fibrillation; Brugada syndrome; progressive and non-progressive heart block; long QT syndrome, sick sinus syndrome; ventricular fibrillation FE: fold enrichment; IA: inverse association; PCOS: polycystic ovarian syndrome; PE: pulmonary embolism; AF: atrial fibrillation

We systematically cross-referenced our results with previously described GWAS associations (via GWAS Catalog^10^ and PhenoScanner^11^) and disease relevance as reported in ClinVar^12^ (**Methods**). In total, we found that 216 of 620 (34.8%) distinct region-disease cluster associations had not previously been reported at *p*<5×10^−8^ (130/445 [29.2%] at *p*<2×10^−9^). Of the 216 distinct loci, 177 (104/130 at *p*<2×10^−9^) were in genes not previously mapped to the respective diseases (**Supplementary Table 3, Figure 1a, Supplementary Figure 1 (interactive)**). Of the novel associations at GWAS significance (*p<*5×10^−8^), roughly one third had MAF<5% in either UKB or FG and 15% had MAF<1% (**Supplementary Table 3**). Importantly, 17% of known (UKB; 19% in FG), but 31% of novel (UKB; 28% in FG) associations had a MAF<5%. Correspondingly, 5% of known (UKB; 6% in FG) and 15% of novel (UKB; 10% in FG) associations had a MAF<1%, highlighting the power gained through our approach especially in the low and rare allele frequency spectrum (**Figure 1e, Supplementary Table 3)**.

Mapping associations to genes, we found the majority of gene loci (81.2% at *p*<5×10^−8^, MHC region counted as one locus) to be associated with a single disease cluster (**Extended Data Figure 6a**). Thirteen loci were associated with ≥ 5 trait clusters (at *p*<5×10^−8^), including well established pleiotropic regions such as the *MHC, APOE, PTPN22, GCKR, SH2B3* and *FUT2* (**Figure 1a**). For instance, in addition to a known association with breast cancer, we found variants in *CHEK2* as associated with the risk of colorectal and thyroid cancers, uterine leiomyoma, benign meningeal tumours and ovarian cysts. Also, in addition to a known association with prostate hyperplasia, we found an *ODF3* missense variant (rs72878024) to be associated with risk of uterine leiomyoma, benign meningeal tumour, lipoma and polyps in the female genital tract (**Supplementary Table 3**).

Harnessing the added power of UKB and FG, we were able to detect GWAS associations for rare variants previously only annotated as causal for single-gene diseases, establishing a disease relevance for these variants at the population level. Of the 534 distinct variants with significant disease associations in our study (*p<*5×10^−8^), 152 had previously been linked to diseases in ClinVar. For 45 of these variants, the associated disease cluster matched with a previously reported phenotype in ClinVar. Notably, only six of these 45 variants (in *GJB2, ABCC6, BRCA1, SERPINA1, FLG*, and *MYOC*) had a previous annotation as either pathogenic or likely pathogenic (**Supplementary Table 7**), with 15 others annotated as benign. Of the novel trait cluster associations, 17 had been reported in ClinVar for the same/similar diseases, with 4 being classified as pathogenic/likely pathogenic and 13 classified either as benign or having “conflicting interpretation of pathogenicity” for the associated trait (**Supplementary Table 3, Supplementary Table 7**). For instance, we found a rare missense variant annotated as showing conflicting pathogenicity in ClinVar in *VWF* (rs1800386:C; Tyr1584Cys; MAF=0.44% [UKB], 0.47% [FG]) to be associated with the risk of von Willebrand disease^12^ (log[OR]=2.09, *p*=8.7×10^−9^); or a missense variant in *SPINK1* (rs17107315:C; Asn34Ser; MAF=1.3% [UKB], 1.6% [FG]) annotated as showing conflicting pathogenicity in ClinVar for chronic pancreatitis to be associated with chronic pancreatitis risk^12^ (log[OR]=1.16, *p*=6.9×10^−25^) and acute pancreatitis risk (log[OR]=0.69, *p*=2.3×10^−18^). These examples highlight that population-scale analyses like ours can help refine pathogenicity assignments through contributing quantitative information on relative disease risks for variant carriers. Likewise, 17 of the 23 genes with highly population-enriched sentinel variants (**Table 1**) were OMIM listed disease genes. Of these, 10 (*CHEK2, DBH, SCL24A5, CFI, FLG, XPA, F10, BRCA1, SCN5A, CACNA1D*) showed associations with conditions identical or related to the respective Mendelian disease, unveiling a relevance of the associated variants on the population level. For instance, we found the missense variant rs77273740 in *DBH* (enriched by >50x in FG), a gene associated with orthostatic hypotension, to be associated with *reduced* risk of hypertension (log[OR]=-0.19, *p*=1.3×10^−23^), whilst an in-frame deletion (rs1250342280) in *CACNA1D* (enriched by 4.3x in UKB), a gene associated with primary aldosteronism, was associated with *increased* risk of hypertension (log[OR]= 0.19, *p*=2.0×10^−8^) (**Table 1**).

### Biomedical insights from coding variant associations

We leveraged the coding variant associations identified in our study to generate biological insights for a range of distinct genes, pathways and diseases and in the following exemplify the broad utility of our resource through a set of selected use cases.

### New roles of coagulation pathway proteins in conferring pulmonary embolism risk

We found known and novel associations with pulmonary embolism (PE) risk, including two rare variant associations (average MAF<1%) in genes encoding components of the coagulation cascade at the convergent common pathway (**Figure 2a**). For instance, we discovered a rare missense mutation in *F10*, enriched by ∼5-fold in FG (rs61753266:A; Glu142Lys; MAF=0.33% [UKB], 1.85% [FG]), to be protective against PE (log[OR]=-0.44, *p*=2.9×10^−9^). This variant has been associated with reduced plasma coagulation factor X (beta=-1.12, *p=*2.0×10^−8^) and factor Xa (beta=-1.54, *p=*7.9×10^−15^) levels previously^13^, as well as clinical factor X deficiency^14^. Deficiencies in coagulation factors, including factor X, are associated with increased bleeding liability and reduced thrombotic risk. In a similar fashion, we found a previously reported venous thromboembolism risk-reducing variant (rs4525:C; His865Arg; MAF=27.2% [UKB], 22.3% [FG]) in *F5* that is also protective for PE (log[OR]=-0.14, *p*=1.2×10^−15^) and associated with reduced plasma F5 levels^15^ (beta=-0.25, *p*=6.0×10^−7^). This variant acts opposite to the well-established risk promoting F5 Leiden missense mutation, which leads to increased resistance to activated protein C cleavage^16^ and thromboembolism liability, thus unravelling that coding variants in *F5* can have opposite effects on PE risk at the population level (**Figure 2b)**. We performed Mendelian randomisation (MR) using rs4525 and rs61753266 as instruments to estimate the relative reduction in PE risk due to reduced F5 (beta_MR_=0.57, *p*=1.0×10^−15^) and F10 levels (F10: beta_MR_=0.40, p=2.9×10^−9^; F10a: beta_MR_=0.28, p=2.9×10^−9^) respectively (**Figure 2b)**. MR results support the expected clinical indication of factor X inhibitors in thromboembolic diseases and the hypothesis that developing drugs inhibiting factor V will also likely be beneficial for PE. We also found a rare variant in fibrinogen (*FGB* rs2227434:T; Pro100Ser; MAF=0.13% [UKB], 0.15% [FG], **Figure 2a, Supplementary Table 3**) that associated with increased PE risk at nominal GWAS significance (log[OR]=1.03, *p*=1.5×10^−8^). Missense mutations in *FGB* have previously been linked to both elevated and reduced fibrinogen levels through GWAS^17,18^, as well as congenital afibrinogenemia^19^.

**Figure 2.**
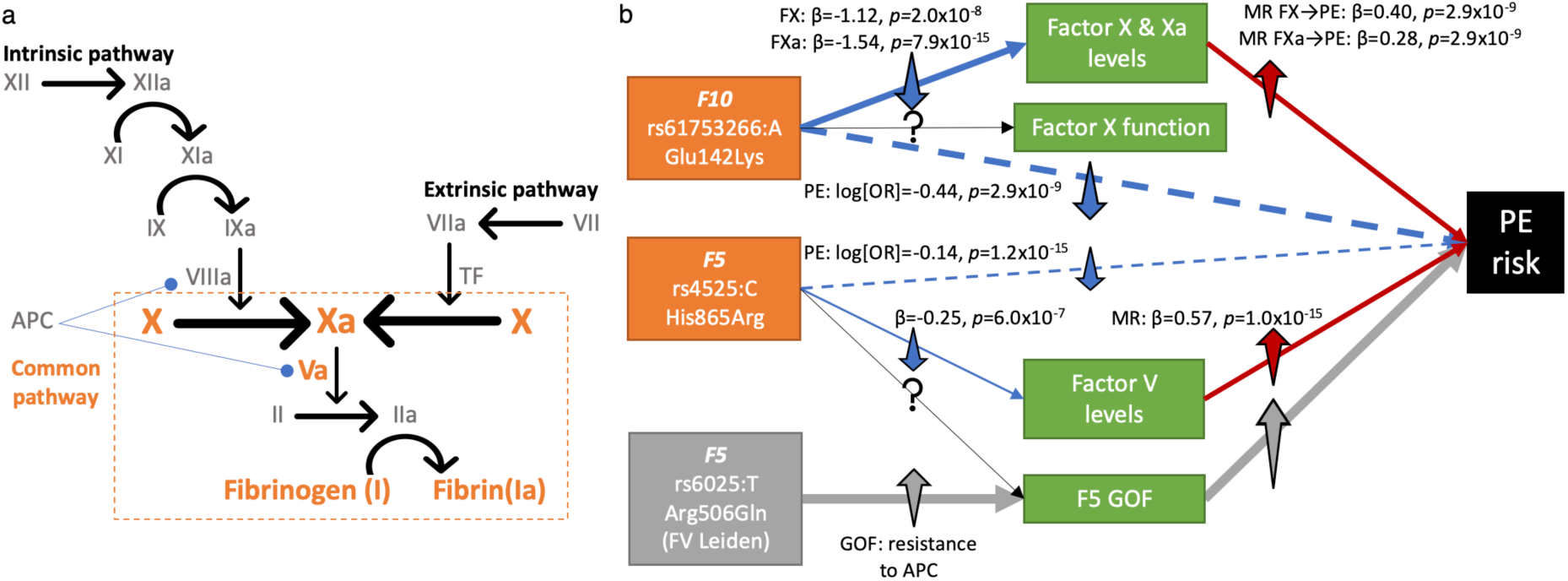
**(a) Simplified diagram of the coagulation cascade**. Factors (in roman numerals, “a” represents activated) with genetic association with PE highlighted in orange. Blue line (round end) indicates inhibitory effect of APC on VIIIa and Va. **(b) Schematic of potential pathway from missense variants in *F5* and *F10* to PE risk**. Factor V Leiden variant had null associations with F5 levels (β _F5 levels_=0.21, *p=*0.091). Dashed blue lines suggest effect of the variants on PE risk which we assume under MR framework acts through factor levels (solid blue lines). Grey box and arrows represent known pathway for Factor V Leiden mutation. *GOF: Gain of function, APC: Activated protein C, MR: Mendelian randomisation, PE: Pulmonary embolism*.

### Rare variant biomarker associations yield insights into disease mechanisms

We interrogated the sentinel variants identified in this study for associations with 117 quantitative biomarkers spanning eight categories in UKB (**Supplementary Table 8**). At a multiple testing adjusted threshold of *p*<1×10^−6^, we found 108 of the biomarkers to be associated with at least one of 417 sentinel variants across 239 regions (**Figure 3a, Supplementary Table 9)**. 47 of the regions were associated with 5 or more biomarker categories (**Extended Data Figure 6b, Supplementary Table 9**), including pleiotropic disease loci such as *MHC, APOE, GCKR, SH2B3, FUT2, MC1R, ABCG5*.

**Figure 3.**
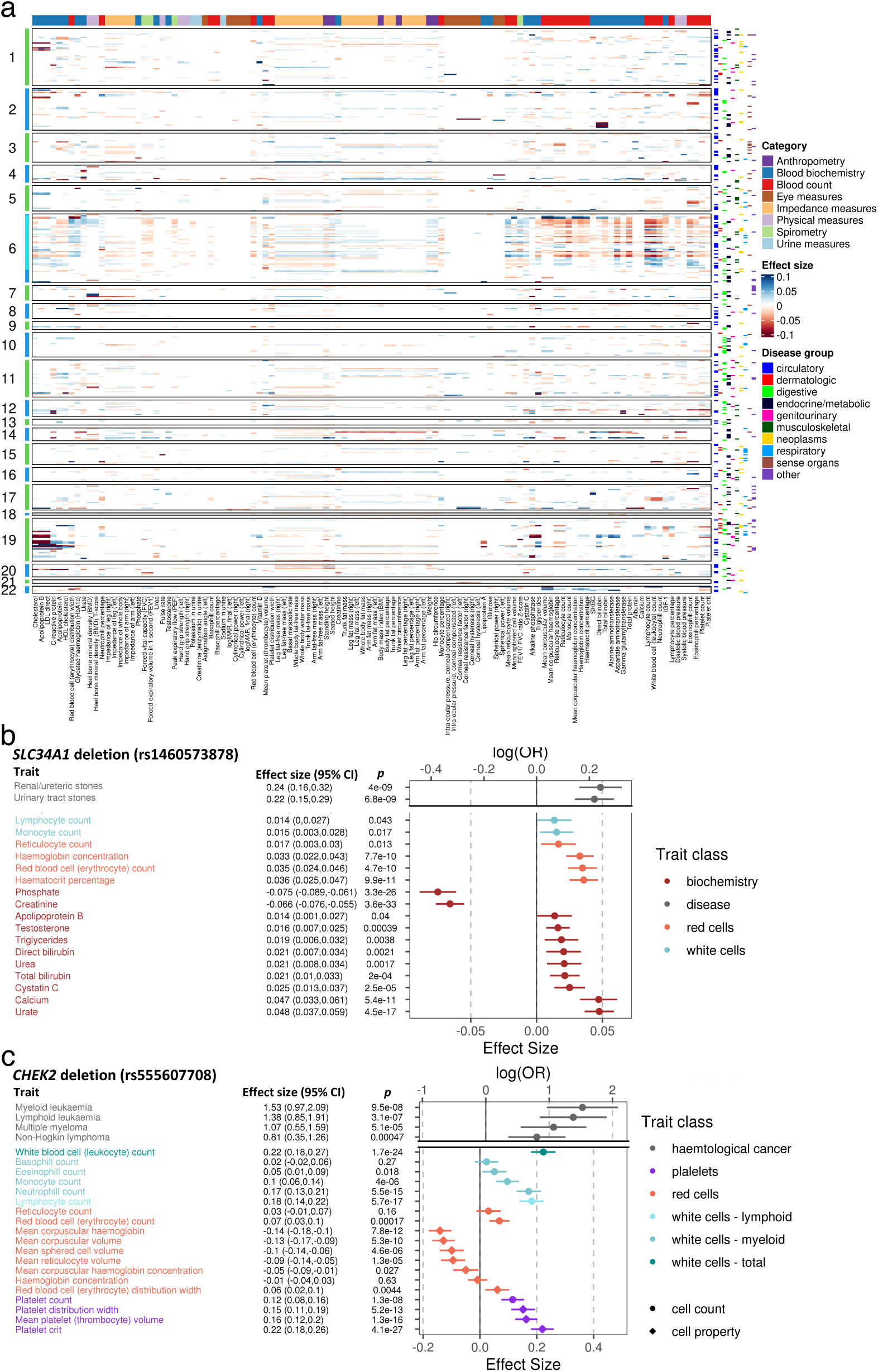
Biomarker associations with sentinel variants. **(a) Heatmap of sentinel associations with biomarkers.** Only significant associations (*p*<10^−6^) displayed. Left side indicates chromosome with cyan indicating MHC region. Right-side: sentinel association with disease by group (colours). Top colours: category of biomarkers. **(b) Forest plot of associations between *SLC34A1* deletion (rs1460573878) with haematological and biochemistry biomarkers**. *P*<0.05 displayed. **(c) Forest plot of associations between *CHEK2* deletion (rs555607708) with haematological biomarkers**.

Many of the newly discovered associations with biomarkers are biologically plausible. For example, a low-frequency missense variant in *ADH1B* (rs1229984:T; Arg48His; MAF=2.2% [UKB], 0.5% [FG]) that is associated with increased enzymatic activity of alcohol dehydrogenase and reduced alcohol tolerance, is also associated with reduced risk of alcohol-related disorders (alcoholic liver disease: log[OR]=-1.08, *p*=1.5×10^−9^; mental and behavioural disorders due to alcohol: log[OR]=-0.82, *p*=1.2×10^−33^) and increased risk of gout (log[OR]=0.39, *p*=3.3×10^−10^). Notably, the alcohol dependence disorder-promoting *ADH1B* allele (C) is also associated with reduced IGF-1 (beta=-0.11, *p*=1.5×10^−51^) and vitamin D levels (beta=-0.049, *p*=2.6×10^−10^), increased levels of liver enzymes (alkaline phosphatase: beta=0.087, *p*=3.1×10^−37^; gamma-glutamyl transferase: beta=0.041, *p*=1.62×10^−9^) and total bilirubin (beta=0.031, *p*=5.1×10^−7^), macrocytosis with increased mean corpuscular volume (beta=0.047, *p*=4.2×10^−12^) and mean corpuscular haemoglobin (beta=0.048, *p*=1.4×10^−12^), as well as reduced erythrocyte count (beta=-0.034, *p*=3.2×10^−8^). The gout risk reducing C allele is associated with reduced urate levels (beta=-0.061, *p*=1.4×10^−21^).

#### A deletion in SLC34A1 is associated with multiple blood and urinary abnormalities

Cross-referencing with biomarkers provided mechanistic insights into novel findings. For instance, we discovered a novel association between a low frequency in-frame deletion in *SLC34A1* (rs1460573878, also known as rs876661296; MAF=2.6% [UKB], 2.7% [FG]; p.Val91_Ala97del) coding for the type II sodium phosphate cotransporter, NPT2a, which is expressed specifically in renal proximal tubular cells, to be associated with increased risk of renal (log[OR]=0.24, *p*=4.0×10^−9^) and urinary tract stones (log[OR]=0.21, *p*=6.8×10^−9^). The deletion has previously been implicated in hypercalciuric renal stones^20,21^ and autosomal recessive idiopathic infantile hypercalcaemia^22^ in family studies. The variant is also associated with increased serum calcium (beta=0.047, *p*=5.4×10^−11^) and reduced phosphate (beta=-0.075, *p*=3.3×10^−26^), consistent with a disrupted function/cell surface expression of the transporter^22^ (**Figure 3b**). We further find associations with increased levels of serum urate (beta=0.048, *p*=4.5×10^−17^), suggesting an increased risk also of uric acid stones. Additionally, we found associations with increased erythrocyte count (beta=0.035, *p*=4.7×10^−10^), haemoglobin concentration (beta=0.033, *p*=7.7×10^−10^) and haematocrit percentage (beta=0.036, *p*=9.9×10^−11^), suggesting increased renal-driven erythropoiesis (**Figure 3b)**. Serum creatinine was not increased in carriers of the deletion (beta=-0.07, *p*=3.6×10^−33^), suggesting renal function is not adversely affected in deletion carriers. Amongst 11,114 renal/ureteric and 13,319 urinary tract stone cases, we identified 735 (renal/ureteric) and 863 (urinary tract) carriers of the deletion who may benefit from clinical interventions targeting NPT2a related pathways and monitoring for deranged biochemical and haematological biomarkers.

#### A CHEK2 deletion is associated with blood cell counts and haematological malignancies

A frameshift deletion in *CHEK2* (rs555607708; MAF=0.64% [FG], 0.24% [UKB]) that increases breast cancer risk has been previously implicated also in myeloproliferative neoplasms through GWAS^23^ and lymphoid leukaemia in a candidate variant study^24^. Consistently, we found nominally significant associations with risks of both, myeloid (log[OR]=1.52, *p*=9.5×10^−8^) and lymphoid (log[OR]=1.38, *p*=3.1×10^−7^) leukaemia, but also multiple myeloma (log[OR]=1.07, *p*=5.1×10^−5^) and non-Hodgkin lymphoma (log[OR]=0.81, *p*=4.7×10^−4^). Association of rs555607708 with clinical haematology traits showed statistically significant associations with increased blood cell counts for both, myeloid (leukocytes, neutrophils, platelets at *p<*1×10^−6^; monocyte and erythrocytes at *p<*1×10^−3^) and lymphoid (lymphocytes, *p*=5.7×10^−17^) lineages (**Figure 3c)**. Furthermore, we found associations with increased mean platelet volume (MPV, *p*=1.3×10^−16^) and platelet distribution width (PDW, *p*=5.2×10^−13^), consistent with increased platelet activation and previous associations of MPV and PDW with chronic myeloid leukaemia^25^. We also found associations with decreased mean corpuscular haemoglobin (*p*=7.8×10^−12^) and mean corpuscular volume (*p*=5.3×10^−10^), suggesting predisposition to haematological cancers by loss-of *CHEK2* function is accompanied by a microcytic red blood cell phenotype (**Figure 3c)**.

### Coding variant associations inform drug discovery and development

We cross-referenced genes with significant coding variant associations with drug targets using the therapeutic targets database^26^. We found 66 genes with trait cluster associations that are the targets of either approved drugs (26 genes) or drugs currently being tested in clinical trials (40 genes), among these, 14 in phase 3 trials (**Supplementary Table 10**). We found a statistical enrichment for significant genes in our study to also be approved drug targets (26/482; compared with a background of 569 approved targets/19,955 genes, OR=1.9, *p*=0.0024), which is in line with previous estimates of a higher success rate for drug targets supported by genetics^27,28^. Sensitivity analyses using more stringent association *p*-value thresholds further increased these probability estimates (*p=*5×10^−9^ [OR 2.3, *p*=0.00070]; *p=*5×10^−10^ [OR 2.5, *p*=0.00037], supporting previous observations that the stronger the genetic association, the higher the likelihood of therapeutic success (**Supplementary Table 11**). In addition to providing further support for well-established drug target associations such as between *PCSK9* loss-of-function and hypercholesterolaemia, or *F10* loss-of-function and venous thromboembolism, we also found an association between a common missense variant (rs231775:G) in *CTLA4* with increased risk of thyrotoxicosis (log[OR]=0.12, *p*=8.5×10^−13^).

Since this variant is also a blood eQTL for decreased *CTLA4* expression^29^ (Z-score=-6.91, *p=*5.0×10^−12^), the association between genetic CTLA4 reduction and thyroid dysfunction might contribute to the adverse event of hyperthyroidism in cancer patients treated with CTLA4 inhibitors^30^.

Genetics can inform drug discovery also on alternative indications for repurposing. For example, *TYK2* inhibitors are being tested in clinical trials for various autoimmune and psoriatic diseases^31^. Consistent with previous GWAS^10^, we found a missense variant in *TYK2* (rs34536443:C) to be associated with reduced risk of rheumatoid arthritis and psoriatic diseases (**Supplementary Table 3**). Our analyses establish this variant to also be associated with sarcoidosis (log[OR]=-0.41, *p*=3.6×10^−8^), proposing sarcoidosis as a new indication for *TYK2* inhibitors. Similarly, while the pleiotropy of *CHEK2* provides support for exploring CHEK2 inhibitors against a broader spectrum of malignancies, our analyses also highlight a risk for potential haematological perturbations upon CHEK2 inhibitor treatment.

### Genetic insights into atrial fibrillation

Atrial fibrillation (AF) GWAS have yielded a sizeable number of loci^32,33^. We chose AF to exemplify how results from our study can further elucidate the genetics and biological basis of one distinct human trait, with a particular emphasis on how our results might help to disambiguate AF loci to causal genes and explain the functional significance of coding variant associations. Indeed, we report several coding variant associations (**Supplementary Table 3**) where prior GWAS ^32,33^ had fallen short to resolve GWAS loci to coding genes and explain disease mechanisms.

#### A binding motif disrupting missense variant reveals a role for METTL11B methylase in AF

The AF GWAS sentinel variant rs72700114 is an intergenic variant located between *METTL11B* and *LINC01142*^32-34^. Our study unveiled a low frequency missense variant in *METTL11B* (rs41272485:G; Ile127Met; MAF=3.9% [UKB], 3.8% [FG]) as associated with increased AF risk (log[OR]=0.14, *p*=4.0×10^−11^). METTL11B is a N-terminal monomethylase that methylates target proteins containing an N-terminal [Ala/Pro/Ser]-Pro-Lys motif^35^. The missense variant Ile127Met (SIFT=0, PolyPhen=1.0) falls within a conserved motif in the enzyme’s S-adenosylmethionine/S-adenosyl-l-homocysteine ligand binding site^36^. *METTL11B* expression is enriched in heart and skeletal muscles with highest expression in heart muscle, in particular cardiomyocytes^37,38^. We scanned protein sequences for a presence of the [Ala/Pro/Ser]-Pro-Lys motif and elevated expression in cardiomyocytes (**Methods, Supplementary Table 12**). We found statistically significant enrichment of genes encoding [Ala/Pro/Ser]-Pro-Lys motif containing proteins amongst genes with elevated expression in cardiomyocytes (OR=1.34, 95% CI=[1.16, 1.54], *p=*3.2×10^−5^), many of which show N-terminal [Ala/Pro/Ser]-Pro-Lys motifs (OR=1.24, 95% CI=[1.06, 1.44], *p=*5.6×10^−3^). This group included several well-established AF genes^39^ such as potassium channels (*KCNA5, KCNE4, KCNN3*), sodium channels (*SCN5A, SCN10A*), *NPPA*, and *TTN*. Our data support *METTL11B* as the causal gene in this GWAS locus and a relevance for N-terminal [Ala/Pro/Ser]-Pro-Lys methylation in cardiomyocytes for AF.

#### Rare variants unveil causal mechanisms in SCN5A-SCN10A and HCN4-REC114 AF loci

Within the *SCN5A-SCN10A* locus, we replicated a common missense variant in *SCN10A* (rs6795970:A; Ala1073Val; MAF=40.0% [UKB], 44.6% [FG]) that was previously described to prolong cardiac conduction^40^. Additionally, we found associations with reduced AF risk (log[OR]=-0.06, *p*=2.1×10^−12^), reduced pulse rate (beta=-0.02, *p*=4.8×10^−18^), and a suggestive signal for increased risk of atrioventricular block (log[OR]=0.10, *p*=1.9×10^−7^). On top of this, we discovered a rare, FG enriched missense variant (rs45620037:A; Thr220Ile; MAF=0.11% [UKB], 0.47% [FG]; SIFT=0.03, PolyPhen=0.96) in *SCN5A*, encoding the cardiac sodium channel Na_V_1.5, as associated with decreased risk of AF (log[OR]=-0.65, *p*=1.3×10^−12^). The variant lies in the S4 voltage sensing segment of the first transmembrane domain of SCN5A^41^ and leads to a substitution of the polar Thr to a non-polar Ile residue, most probably causing loss of function and electrophysiological changes^42,43^. The Thr220Ile variant has been associated with dilated cardiomyopathy^44^ and conduction defects including sick sinus syndrome and atrial standstill^45^ in family studies with bradycardic changes. Consistently, we found a nominal association with reduced pulse rate (beta=-0.078, *p=*0.023), suggesting that protective effects of the variant will be most beneficial for the common tachycardic form of AF through reducing pulse rate. Notably, our results identify both, *SCN5A* and *SCN10A* as likely causal genes at this AF locus.

Another AF GWAS locus is tagged by the common intergenic sentinel variant rs74022964 between *HCN4* and *REC114*^32,33^. We identified a rare, FG enriched variant in *HCN4* (rs151004999:T; Asp364Asn; MAF=0.045% [UKB], 0.17% [FG]; SIFT=0.05, PolyPhen=0.41) as associated with increased AF risk (log[OR]=0.72, *p*=2.8×10^−8^). HCN4 is a hyperpolarization-activated ion channel contributing to cardiac pacemaker (funny) currents (I_f_). Mutations in *HCN4* have been associated with familial bradycardia (also known as sick sinus syndrome 2) and Brugada syndrome 8 in family studies^46^. Consistently, in our study we also found an association with decreased heart rate (beta=-0.49, *p*=3.8×10^−21^).

#### Genetic variants underlying AF risk differentially modulate pulse rate

To further evaluate the hypothesis that distinct genetic mechanisms underlying AF risk inversely modulate pulse rate, we applied clustered Mendelian randomization (MR-Clust)^47^ with a slight modification to the mixture-model to better accommodate rare-variants. Specifically, we related expectation maximization clustering of AF associated variants with homogenous directional effects on pulse rate (**Methods**). Among the AF sentinel variants from our coding variant association analyses, we found the two above described clusters of variants in *SCN10A* (rs6795970) and *HCN4* (rs151004999), suggestive of two genetic components of AF that can increase and decrease pulse rate, respectively (**Figure 4a, left**). Identifying components of AF with diverging directionality on pulse rate is not surprising given clinically AF can be driven by both tachycardia and bradycardia through distinct mechanisms^48^. As sensitivity analyses, we used sentinel variants from a recent AF GWAS^32^ where the candidate gene sets overlapped those in our study and found concordant patterns (**Figure 4a, left-middle, Supplementary Table 13**). We also included all sentinel variants in the AF GWAS loci^32^ and found additional clusters with differing impact on pulse rate (**Figure 4a, right-middle**). Conversely, as expected, permuting pulse rate led to no clustering and null associations between AF and pulse (**Figure 4a, right**). Expectedly, within the AF GWAS loci^32^ the two rare missense alleles in *HCN4* (rs151004999:T, log[OR]=0.72) and *SCN5A* (rs45620037:A, log[OR]=-0.65) identified in our study had much larger effect sizes on AF risk than the respective non-coding sentinel GWAS variants (rs74022964:T [*HCN4* locus], log[OR]=0.12; rs6790396:C [*SCN5A, SCN10A* locus], log[OR]=-0.058) (**Figure 4a**).

**Figure 4.**
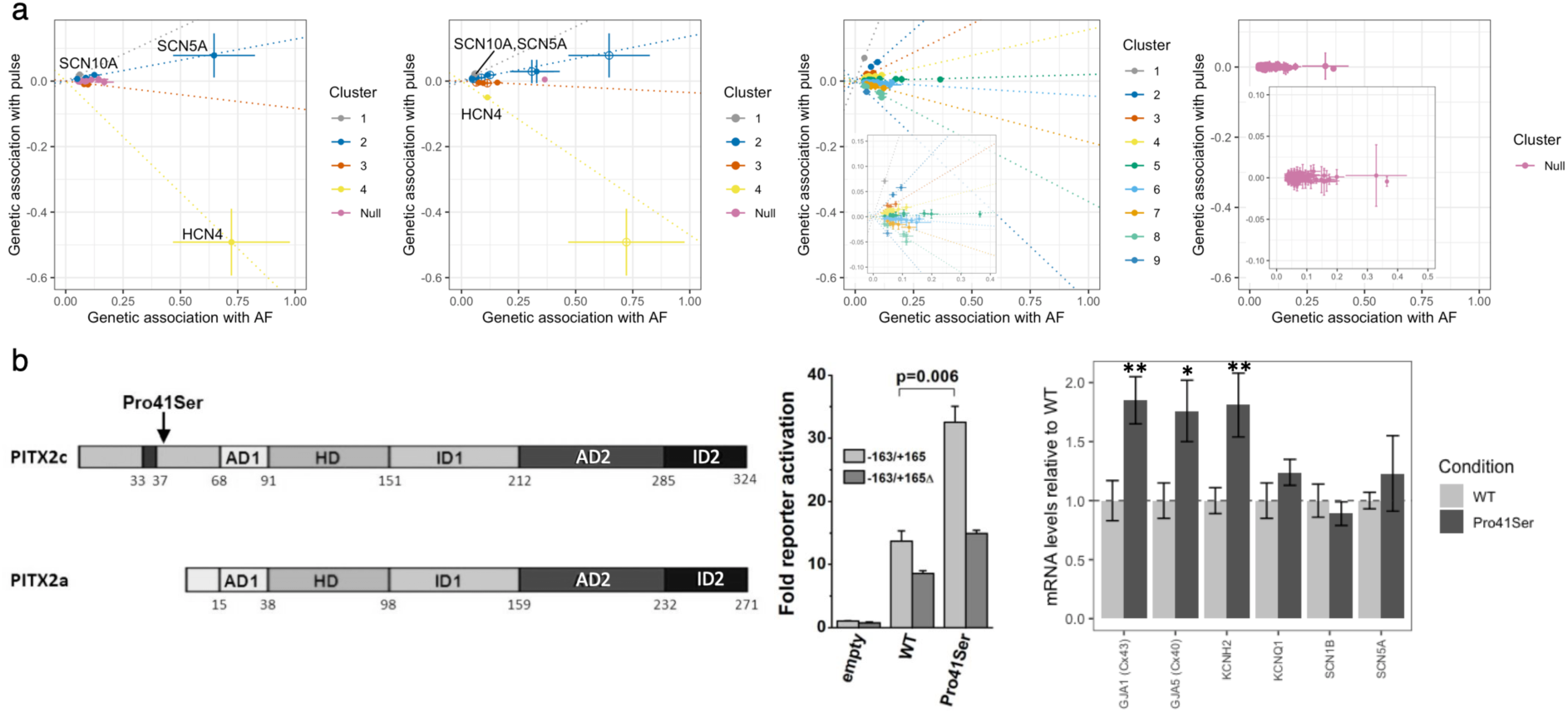
Genetic and functional insights into AF. **(a) Clustered MR plot of AF loci on pulse rate**. Only variants with cluster inclusion probability>0.7 are included. Left to right: CWAS loci (sentinels), Overlapping CWAS and AF GWAS loci, All AF GWAS loci from Nielsen *et al*. (with zoomed inset), All AF GWAS loci with permuted pulse (null, with zoomed inset). (**b) Functional effect of PITX2c Pro41Ser variant (rs143452464) in vitro. Left**: schematic of the location of the Pro41Ser variant in PITX2c as compared to the PITX2a splicing alternative. AD1: common sequence, HD: homeodomain, ID1: transcriptional inhibitory domain 1, AD2: second common sequence, ID2: transcriptional inhibitory domain 2. Pro41Ser is within the N-terminal domain (grey), near to the 5-AA sequence (33 to 37 red, LAMAS) important for transcriptional activity of the N-terminal of PITX2c. **Middle**: **Reporter gene assays in TM-1 cells**. Luciferase values from activation of the SLC13A3-reporter plasmids (n=3) were normalized to β-galactosidase (expressed from the transfection control plasmid), relative to the ratio for empty expression vector plus non-deleted SLC13A3 reporter (“-163/+165”). The reporter plasmid designated as “-163/+165Δ” contains a deletion of 8bp corresponding to the predicted PITX2 binding site. **Right: qRT-PCR analysis of HL-1 cells transfected with PITX2c recombinant plasmids**. Effect of Pro41Ser PITX2c variant expression on Cx40, Cx43, KCNQ1, KCNH2, SCN1B and SCN5A. ^*^*p*<0.05, ^**^*p*<0.01.

#### PITX2c Pro41Ser increases AF risk through a gain-of-function mechanism

Lastly, we found a rare missense variant in *PITX2* as associated with increased risk of AF (log[OR]=0.38, *p*=1.1×10^−9^). This variant is enriched nearly 50-fold in FG (rs143452464:A; Pro41Ser; MAF=0.023% [UKB], 1.1% [FG]) and was independently identified in a French family with AF (**Supplementary Information, Supplementary Figure Pro41Ser 1**), whilst GWAS had linked intergenic variants between *PITX2* and *FAM241A* to AF risk. PITX2 is a bicoid type homeobox transcription factor previously assumed to play a role in cardiac rhythm control^49^. The Pro41Ser variant lies in the N-terminal domain that is only present in the PITX2c isoform expressed in cardiac muscle (**Figure 4b left**). We performed reporter assays comparing the ability of Xpress-*PITX2c* constructs to transactivate a luciferase reporter plasmid containing a putative PITX2c binding element (**Supplementary Information)**. A construct containing the Pro41Ser variant showed a ∼2.4-fold higher activation of the reporter than the wild-type construct (*p*=0.006, **Figure 4b middle**). This effect was abrogated upon deletion of the putative PITX2c binding site (**Figure 4b middle**; see also **Supplementary Information, Supplementary Figure Pro41Ser 2** and **Supplementary Figure Pro41Ser 3)**. In cultured cardiac muscle HL-1 cells, Pro41Ser increased the transcription of several presumed PITX2c target genes, specifically *GJA1* (*Cx40*, 1.76-fold, *p=*0.012), *GJA5* (*Cx43*, 1.85-fold, *p=*0.005) and *KCNH2* (1.81-fold, *p=*0.009), while the transcription of other selected genes with putative roles in AF was not substantially altered (**Figure 4b right, Supplementary Table 14**, see also **Supplementary Information**). Together, these results are consistent with a putative gain-of-function mechanism of Pro41Ser on PITX2c transactivation potential and AF risk.

## Discussion

Here, we have conducted the to date largest association study of protein-coding genetic variants against hundreds of disease endpoints ascertained from two massive population biobanks, UK Biobank and FinnGen. We report novel disease associations, most notably in the rare and low allelic frequency spectrum, replicate and assign putative causalities to many previously reported GWAS associations, and leverage the insights gained to elucidate disease mechanisms. In addition to a substantial gain in power over previous studies, our analyses benefit from replication between two population cohorts, increasing the robustness of our findings and setting the stage for future similar studies in ethnically more diverse populations.

Importantly, our study identifies both, pathogenic variants residing in monogenic disease genes to impact the risk for related complex conditions, as well as new, likely causal sentinel variants within GWAS loci in genes with known and novel biological roles in the respective GWAS trait. With this, our study is one of the first to help bridge the gap between common and rare disease genetics across a broad range of conditions and provides support for the hypothesis that the genetic architecture of many diseases is continuous^1^. As reflected in a recent schizophrenia study^50^, GWAS tend to identify association signals primarily for variants with MAF>2%, while most variants identified through exome sequencing are ultra-rare (MAF<0.01%). Of the 975 associations identified in our study, 145 are driven by unique variants in the yet little interrogated rare and low allelic frequency spectrum that is hypothesized to contribute substantially to the “missing heritability” of many human diseases^51^.

Our approach benefits considerably from the Finnish genetic background where, consistent with previous observations^6-8^, certain alleles are stochastically enriched to unusually high allele frequencies, at times exceeding population frequencies in the UK Biobank by >50-fold (such as for instance the *PITX2*-Pro41Ser variant). Our theoretical and empirical results suggest the increasing utility of enriched variants for identifying associations quantitatively towards lower allelic frequencies. Notably, we identify the most prominent relative power gain in the rarest variant frequency spectrum, highlighting a role for sequencing studies and integrating additional population cohorts with enriched variants for identifying novel disease associations at scale. In our study, we identify several alleles with comparatively high effect sizes and a prevalence in the population that warrants follow-up, both experimentally as well as potentially directly in clinical settings to help improve disease outcomes. For instance, our data propose that 6.5% of UKB and FG participants with kidney or urinary tract stones, conditions debilitating >15% of men and 5% of women by 70 years of age^52^, carry a deletion in *SLC34A1*. Monitoring patients for the clinical biomarkers identified here as associated with this deletion might help to differentiate aetiologies and guide individualized treatments. Likewise, coding variant associations identified in our study may serve as an attractive source to generate hypotheses for drug discovery programs. Our results support previous studies^27,28^ that drug targets supported by human genetics have an increased likelihood of success, which can be considered particularly high when the genetic effect on a drug target closely mimics that of a pharmacological intervention^53^.

We demonstrate the broad utility of our results through numerous examples. For instance, in the case of AF we show how coding variants can help disambiguate GWAS loci to likely causal genes and in some cases predict specific changes in a protein’s function; how integration of genetics with intermediate traits (such as slow versus fast heart rate) can unravel different biological mechanisms underlying a disease entity; or how a putative function of sentinel coding variants can be further validated through experiments. Our examples highlight that the step from association to biological insight may be considerably shorter for coding variant association studies than it has traditionally been for GWAS.

The results of our study foreshadow the discovery of many additional coding and non-coding associations from cross-biobank analyses at even larger sample sizes. With the continued growth of population biobanks with comprehensive health data also in non-European populations, the emergence of more and more cost-effective technologies for sequencing and genotyping, and computational advances to analyse genetic and non-genetic data at scale, future studies will be able to assess the genetic contribution to health and disease at even finer resolution.

## Methods

### Samples and participants

UK biobank (UKB) is a UK population study of approximately 500,000 participants aged 40-69 years at recruitment^2^. Participant data include genomic, electronic health record linkage, blood, urine and infection biomarkers, physical and anthropometric measurements, imaging data and various other intermediate phenotypes that are constantly being updated. Further details are available at https://biobank.ndph.ox.ac.uk/showcase/. Analyses in this study were conducted under UK Biobank Approved Project number 26041.

FinnGen (FG) is a public-private partnership project combining electronic health record and registry data from six regional and three Finnish biobanks. Participant data include genomics and health records linked to disease endpoints. Further details are available at https://www.finngen.fi/. More details on FG and ethics protocols are provided in **Supplementary Information**. We used data from FG participants with completed genetic measurements (R5 data release) and imputation (R6 data release). FinnGen participants provided informed consent for biobank research. Recruitment protocols followed the biobank protocols approved by Fimea, the National Supervisory Authority for Welfare and Health. The Coordinating Ethics Committee of the Hospital District of Helsinki and Uusimaa (HUS) approved the FinnGen study protocol Nr HUS/990/2017. The FinnGen study is approved by Finnish Institute for Health and Welfare.

### Disease phenotypes

FG phenotypes were automatically mapped to those used in the Pan UKBB (https://pan.ukbb.broadinstitute.org/) project. Pan UKBB phenotypes are a combination of Phecodes^54^ and ICD10 codes. Phecodes were translated to ICD10 (https://phewascatalog.org/phecodes_icd10, v.2.1) and mapping was based on ICD-10 definitions for FG endpoints obtained from cause of death, hospital discharge and cancer registries. For disease definition consistency, we reproduced the same Phecode maps using the same ICD-10 definitions in UKB. In particular, we expertly curated 15 neurological phenotypes using ICD10 codes. We retained phenotypes where the similarity score (Jaccard index: ICD10_FG_ ∩ ICD10_UKB_ / ICD10_FG_ ∪ ICD10_UKB_) was >0.7 and additionally excluded spontaneous deliveries and abortions.

Phecodes and ICD10 coded phenotypes were first mapped to unified disease names and disease groups using mappings from Phecode, “PheWAS” and “icd” R packages followed by manual curation of unmapped traits and diseases groups, mismatched and duplicate entries. Disease endpoints were mapped to Experimental Factor Ontology (EFO) terms using mappings from EMBL-EBI and Open Targets based on exact disease entry matches followed by manual curation of unmapped traits.

Disease trait clusters were determined through first calculating the phenotypic similarity via the cosine similarity, then determining clusters via hierarchical clustering on the distance matrix (1-similarity) using the Ward algorithm and cutting the hierarchical tree, after inspection, at height 0.8 to provide the most semantically meaningful clusters.

### Genetic data processing

#### UKB genetic QC

UKB genotyping and imputation were performed as described previously^2^. WES data for UKB participants were generated at the Regeneron Genetics Center (RGC) as part of a collaboration between AbbVie, Alnylam Pharmaceuticals, AstraZeneca, Biogen, Bristol-Myers Squibb, Pfizer, Regeneron and Takeda with the UK Biobank. WES data were processed using the RGC SBP pipeline as described in ^3,55^. RGC generated a QC-passing “Goldilocks” set of genetic variants from a total of 454,803 sequenced UK Biobank participants for analysis. Additional QC were performed prior to association analyses as detailed below.

#### FG genetic QC

Samples were genotyped with Illumina (Illumina Inc., San Diego, CA, USA) and Affymetrix arrays (Thermo Fisher Scientific, Santa Clara, CA, USA). Genotype calls were made with GenCall and zCall algorithms for Illumina and AxiomGT1 algorithm for Affymetrix data. Sample, genotyping as well as imputation procedures and QC are detailed in **Supplementary Information**.

#### Coding variant selection

GnomAD v.2.0 variant annotations were used for FinnGen variants^56^. The following gnomAD annotation categories are included: predicted loss-of-function (pLoF), low-confidence loss-of-function (LC), in-frame indel, missense, start lost, stop lost, stop gained. Variants have been filtered to imputation INFO score > 0.6. Additional variant annotations were performed using variant effect predictor (VEP)^57^ with SIFT and PolyPhen scores averaged across the canonical annotations.

### Disease endpoint association analyses

For optimized meta-analyses with FG, analyses in UKB were performed in the subset of exome-sequence UKB participants with white European ancestry for consistency with FG (n=392,814). We used REGENIE v1.0.6.7 for association analyses via a two-step procedure as detailed in^58^. In brief, the first step fits a whole genome regression model for individual trait predictions based on genetic data using the leave one chromosome out (LOCO) scheme. We used a set of high-quality genotyped variants: minor allele frequency (MAF)>5%, minor allele count (MAC)>100, genotyping rate >99%, Hardy-Weinberg equilibrium (HWE) test *p*>10^−15^, <5% missingness and linkage-disequilibrium (LD) pruning (1000 variant windows, 100 sliding windows and r^2^<0.8). Traits where the step 1 regression failed to converge due to case imbalances were subsequently excluded from subsequent analyses. The LOCO phenotypic predictions were used as offsets in step 2 which performs variant association analyses using the approximate Firth regression detailed in^58^ when the *p*-value from the standard logistic regression score test is below 0.01. Standard errors (SEs) were computed from the effect size estimate and the likelihood ratio test p-value. To avoid issues related to severe case imbalance and extremely rare variants, we limited association test to phenotypes with >100 cases and for variants with MAC 5 in total samples and MAC 3 in cases and controls. The number of variants used for analyses varies for different diseases as a result of the MAC cut-off for different disease prevalence. The association models in both steps also included the following covariates: age, age^2^, sex, age^*^sex, age^2*^sex, first 10 genetic principle components (PCs).

Association analyses in FG were performed using mixed model logistic regression method SAIGE v0.39^59^. Age, sex, 10 PCs and genotyping batches were used as covariates. For null model computation for each endpoint each genotyping batch was included as a covariate for an endpoint if there were at least 10 cases and 10 controls in that batch to avoid convergence issues. One genotyping batch need be excluded from covariates to not have them saturated. We excluded Thermo Fisher batch 16 as it was not enriched for any particular endpoints. For calculating the genetic relationship matrix, only variants imputed with an INFO score >0.95 in all batches were used. Variants with >3% missing genotypes were excluded as well as variants with MAF<1%. The remaining variants were LD pruned with a 1Mb window and r^2^ threshold of 0.1. This resulted in a set of 59,037 well-imputed not rare variants for GRM calculation. SAIGE options for null computation were: “LOCO=false, numMarkers=30, traceCVcutoff=0.0025, ratioCVcutoff=0.001”. Association tests were performed phenotypes with case counts >100 and for variants with minimum allele count of 3 and imputation info >0.6 were used.

We additionally performed sex-specific associations for a subset of gender-specific diseases (60 female diseases and in 50 disease clusters, 14 male diseases and in 13 disease clusters) in both FG and UKB using the same approach without inclusion of sex-related covariates (**Supplementary Table 2**)

We performed fixed-effect inverse-variance meta-analysis combining summary effect sizes and standard errors for overlapping variants with matched alleles across FG and UKB using METAL^60^.

### Definition and refinement of significant regions

To define significance, we used a combination of (1) multiple testing corrected threshold of *p*<2×10^−9^, 0.05/(∼26.8×10^6^) [sum (mean number of variants tested per disease cluster)], to account for the fact that some traits are highly correlated disease subtypes, (2) concordant direction of effect between UKB and FG associations, and (3) *p*<0.05 in both UKB and FG.

We defined independent trait associations through LD-based (r^2^=0.1) clumping ±500Kb around the lead variants using PLINK^61^, excluding the HLA region (chr6:25.5-34.0Mb) which is treated as one region due to complex and extensive LD patterns. We then merged overlapping independent regions (±500Kb) and further restricted each independent variant (r^2^=0.1) to the most significant sentinel variant for each unique gene. For defining region associations across traits, we merged overlapping independent regions for each individual trait.

### Cross reference with known genetic associations

We cross-referenced the sentinel variants and their proxies (r^2^>0.2) for significant associations (*p*<5×10^−8^) of mapped Experimental Factor Ontology (EFO) terms and their descendants in GWAS Catalog^10^ and PhenoScanner^11^. To be more conservative with reporting of novel associations, we also considered whether the most-severe associated gene in our analyses were reported in GWAS Catalog and PhenoScanner. In addition, we also queried our sentinel variants in ClinVar^12^ to define known associations with rarer genetic diseases and further manually curated novel associations for previous genome-wide significant (*p*<5×10^−8^) associations.

### Biomarker associations of lead variants

For the lead sentinel variants, we performed association analyses using the two-step REGENIE approach described above with 117 biomarkers including anthropometric traits, physical measurements, clinical haematology measurements, blood and urine biomarkers available in UKB (detailed in **Supplementary Table 8**).

### Drug target mapping and enrichment

We mapped the annotated gene for each sentinel variant to drugs using the therapeutic target database (TTD)^26^. We retained only drugs which have been approved or are in clinical trial stages. For enrichment analysis of approved drugs with genetic associations, we used Fisher’s exact test on the proportion of significant genes targeted by approved drug against a background of all approved drugs in TTD^26^ (n=595) and 20,437 protein coding genes from Ensembl annotations^62^.

### Mendelian randomization (MR) analyses

#### F5 and F10 effect on pulmonary embolism (PE) risk

The missense variants rs4525 and rs61753266 in F5 and F10 genes were taken as genetic instruments for MR analyses. To assess potential that each factor level is causally associated with PE we employed two-sample MR using summary statistics, with effect of the variants on their respective factor levels obtained from previous large scale (protein quantitative trait loci) pQTL studies^13,15^. Let *β*_*XY*_ denote the estimated causal effect of a factor level on PE risk and *β*_*X*_, *β*_*Y*_ be the genetic association with a factor level (FV, FX or FXa) and PE risk respectively. Then, the MR ratio-estimate of *β*_*XY*_ is given by:

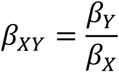

where the corresponding standard error (*β*_*XY*_), computed to leading order, is:

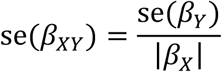

#### Clustered MR

To assess evidence of several distinct causal mechanisms by which atrial fibrillation (AF) may influence pulse rate (PR) we used MR-Clust^47^. In brief, MR-Clust is a purpose-built clustering algorithm for use in univariate MR analyses. It extends the typical MR assumption that a risk factor can influence an outcome via a single causal mechanism^63^ to a framework that allows one or more mechanisms to be detected. When a risk-factor affects an outcome via several mechanisms, the set of two-stage ratio-estimates can be divided into clusters, such that variants within each cluster have similar ratio-estimates. As shown in^47^, two or more variants are members of the same cluster if and only if they affect the outcome via the same distinct causal pathway. Moreover, the estimated causal effect from a cluster is proportional to the total causal effect of the mechanism on the outcome. We included variants within clusters where the probability of inclusion >0.7. We used MR-Clust algorithm allowing for singletons/outlier variants to be identified as their own “clusters” to reflect the large but biologically plausible effect sizes seen with rare and low frequency variants.

### Bioinformatic analyses of motif and expression for *METTL11B*

We searched [Ala/Pro/Ser]-Pro-Lys motif containing proteins using the “peptide search” function on UniProt^64^, filtering for reviewed Swiss-Prot proteins and proteins listed in Human Protein Atlas (HPA)^38^ (n=7,656). We obtained genes with elevated expression in cardiomyocytes (n=880) from HPA based on the criteria: “cell_type_category_rna: cardiomyocytes; cell type enriched, group enriched, cell type enhanced” as defined by HPA in (https://www.proteinatlas.org/humanproteome/celltype/Muscle+cells#cardiomyocytes [accessed 20/03/2021]) with filtering for those with valid UniProt IDs (Swiss-Prot, n=863). Enrichment test was performed using Fisher’s exact test. Additionally, we performed enrichment analyses using any Ala/Pro/Ser]-Pro-Lys motif positioned within the N-terminal half of the protein (n=4,786).

## Supporting information

Supplementary Figure 2c (interactive)

Supplementary Figure 2b (interactive)

Supplementary Figure 2a (interactive)

Supplementary Figure 1 (interactive)

## Data Availability

All data produced in the present study are available upon reasonable request to the authors.

## Acknowledgements

We thank all the participants, contributors and researchers of UK Biobank and FinnGen (and its participating biobanks) for making data available for this study. We thank the UK Biobank Exome Sequencing Consortium (AbbVie, Alnylam Pharmaceuticals, AstraZeneca, Biogen, Bristol-Myers Squibb, Pfizer, Regeneron and Takeda) for generation the whole exome sequencing data and Regeneron Genetics Centre for initial quality control of the exome sequencing data. The FinnGen project is funded by two grants from Business Finland (HUS 4685/31/2016 and UH 4386/31/2016) and the following industry partners: AbbVie Inc., AstraZeneca UK Ltd, Biogen MA Inc., Celgene Corporation, Celgene International II Sàrl, Genentech Inc., Merck Sharp & Dohme Corp, Pfizer Inc., GlaxoSmithKline Intellectual Property Development Ltd., Sanofi US Services Inc., Maze Therapeutics Inc., Janssen Biotech Inc, and Novartis AG. We thank Susanna Lemmelä for her contribution to FinnGen data curation. We further thank Yi-Qing Yang, Tim Footz, Michael Walter, Amelia Aránega, Francisco Hernández-Torres, Elodie Morel and Gilles Millat for their contributions to the functional characterisation of PITX2c. PITX2 functional work was supported in part by grants from the National Natural Science Fund of China (81070153), the Personnel Development Foundation of Shanghai, China (2010019), and the Key Program of Basic Research of Shanghai, China (10JC1414002), and by the Canadian Institutes of Health Research (grants MOP-111072 and MOP-130373 to Mohamed Chahine). Asma Mechakra was supported by a bursary of the French Ministry of Research and Technology (MRT).

## Author contributions

Conceptualization and experimental design: B.B.S., H.R.; methodology: B.B.S., H.R., C.N.F., C.C., M.J.D.; analysis: B.B.S., M.I.K., C.N.F., A.M., C.C., E.M., J.B.W., Biogen Biobank Team; experimental work: A.M., G.C., M.C., P.C.; FinnGen protocols and analysis: M.I.K., A.P., M.J.D., FinnGen; writing: B.B.S., H.R.; all authors critically reviewed the manuscript.

