## Supplementary Information for "Genetic associations of protein-coding variants in human disease"

#### Supplementary Methods: FinnGen genetic QC details

Samples were genotyped with Illumina (Illumina Inc., San Diego, CA, USA) and Affymetrix arrays (Thermo Fisher Scientific, Santa Clara, CA, USA). Genotype calls were made with GenCall and zCall algorithms for Illumina and AxiomGT1 algorithm for Affymetrix data. Chip genotyping data produced with previous chip platforms and reference genome builds were lifted over to build version 38 (GRCh38/hg38) following the protocol described here: dx.doi.org/10.17504/protocols.io.nqtddwn.

##### Sample and initial variant QC

Individuals with ambiguous sex, high genotype missingness (>5%), excess heterozygosity (±4 SD) and non-Finnish ancestry were removed. In variant-wise quality control variants with high missingness (>2%), low Hardy-Weinberg equilibrium (HWE) p-value ($<1x10^{-6}$) and minor allele count, MAC<3 were removed. Chip genotyped samples were pre-phased with Eagle 2.3.5 (https://data.broadinstitute.org/alkesgroup/Eagle/) with the default parameters, except the number of conditioning haplotypes was set to 20,000.

High-coverage (25-30x) WGS data (N= 3,775) used to develop the SISu v3 reference panel were generated at the Broad Institute of MIT and Harvard and at the McDonnell Genome Institute at Washington University; and jointly processed at the Broad Institute. Variant callset was produced with GATK HaplotypeCaller algorithm by following GATK best-practices for variant calling.

##### Additional genotyping QC

Batchwise filters:

1) Not passing Axiom best practices and not recommended by Thermo Fisher Finngen team
2) Proportion of samples missing variant (F_MISS) > 0.02.
3) Hardy-Weinberg equilibrium test p-value ≤ 0.000001.
4) Genotype duplicates marker discrepancy > 2.
5) Mendelian discrepancy in known trios > 2.
6) Allele frequency differences to whole genome reference (SISu v3 imputation panel) and exome datasets (gnomAD exome version 2.1 (Finnish participants) and Finnish exons me collection) log2 fold change ±5 or allele frequency difference 0.1.

7) Variants showing significant differences to population reference panel in GWAS analysis (Sisu v3 imputation panel) p-value < dynamic p-value limit ($5x10^{-8}$/lambda3) (PLINK2, glm firth-fallback).
8) Common variants (AF > 0.05) that are not in reference panel WGS or exon data.

Batches were merged and variants were further filtered:
1) Variant call rate ≥ 80%
2) Hardy-Weinberg Equilibrium test p-value ≤ 0.000001
3) Variant call rate 97.8 % for variants that cannot be validated with allele frequency comparison to reference panel
4) Allele frequency differences to whole genome reference (SISu v3 imputation panel) and exome datasets (gnomAD exome 2.1 panel (Finnish participants) and Finnish exome collection) log2 fold change ±5 or allele frequency difference 0.1.

**Imputation QC**

Genotype-, sample- and variant-wise QC was applied in an iterative manner by using the Hail framework (https://github.com/hail-is/hail) v0.1 and the resulting high-quality WGS data for 3,775 individuals were phased with Eagle 2.3.5 as described above. Genotype imputation was carried out by using the population specific SISu v3 imputation reference panel with Beagle 4.1 (version 08Jun17.d8b, https://faculty.washington.edu/browning/beagle/b4_1.html) as described in the following protocol: dx.doi.org/10.17504/protocols.io.nmndc5e. Post-imputation quality-control involved non-reference concordance analyses, checking expected conformity of the imputation INFO-values distribution, MAF differences between the target dataset and the imputation reference panel and checking chromosomal continuity of the imputed genotype calls.

#### Supplementary Methods: Theoretical description and simulation of the impact of MAF enrichment on inverse-variance weighted (IVW) meta-analysis Z-scores

Here we provide the details of theoretical frameworks and empirical simulations relating MAF enrichment and IVW meta-analysis Z-scores and the interpretation of the results.

Let the parameter $b_i$ denote the effect of genotype $g_i$ on disease endpoint $Y_i$ in the $i$th study, for a total of $i = 1, 2, \dots, m : m \geq 2$ studies. The meta-analysis inverse variance weighted Z-score, which aggregates information across all $m$ studies, is given by

$$Z_{ivw} = \frac{\sum_{i=1}^{m} Z_i / \hat{\sigma}_{b_i}}{\sqrt{\sum_{i=1}^{m} 1 / \hat{\sigma}_{b_i}^2}},$$

where $Z_i$ is the Z-score from the $i$th study, i.e.,

$$Z_i = \frac{\hat{b}_i}{\hat{\sigma}_{b_i}},$$

with $\hat{b}_i$ denoting the sample-based estimate of $b_i$ and $\hat{\sigma}_{b_i}$ being its corresponding standard error. We re-write the above to illustrate the impact of integrating additional independent study information relative to a reference study, which we arbitrarily take to be study $i = 1$. That is, we write $Z_{ivw}$ as:

$$Z_{ivw} = Z_1 \left( \frac{1 + \sum_{i=2}^{m} \frac{\sigma_1 Z_i}{\sigma_i Z_1}}{\sqrt{1 + \sum_{i=2}^{m} \sigma_1^2 / \sigma_i^2}} \right)$$

$$= Z_1 * \alpha$$

$$\Rightarrow \quad \alpha = \frac{Z_{ivw}}{Z_1}.$$

(1)

Hence, $\alpha$ denotes the increase/decrease in Z-score computed on aggregating results across $m$ studies relative to the reference study. When $\alpha > 1$ the aggregated $Z_{ivw}$ score is larger than the reference Z-score ($Z_1$) - a scenario we refer to as 'IVW uplift'. Our goal is to assess changes in $\alpha$ as a function of MAF and MAF-enrichment between $m = 2$ studies. To fix ideas, we are interested in assessing uplift when (i) the reference study is the largest study and (ii) MAF is enriched in the smaller study, i.e., study 2. Hence, $N_1 \geq N_2$ and $MAF_2 \geq MAF_1$. The scenario in which the reference study has larger sample size and also enriched $MAF$, i.e., $N_1 \geq N_2$ and $MAF_1 \geq MAF_2$, is considered later.

**Theoretical description of uplift $\alpha$**

In this section we present a formula which relates uplift $\alpha$ to the core parameters underpinning computation of the test statistic $Z_{ivw}$, i.e., $MAF_i$, disease prevalence ($\pi_i$) and sample size ($N_i$), for each of the $i \in \{1, 2\}$ studies. In doing so, we will (a) illustrate complexity in the relationship between $\alpha$ and the core parameters and (b) highlight the expected utility of a MAF-enriched study design, in particular how the probability of detecting novel (rare variant) associations increases as a function of enrichment.

Let the probability of disease status follow a logistic model, i.e.,

$$\pi_{y_i|g} = P(Y_i = 1 \mid g) = \frac{e^{(a_i+b_i g)}}{1 + e^{(a_i+b_i g)}},$$

(2)

where $g$ denotes a genotype putatively associated with disease risk, $a_i$ is the population baseline effect and $b_i$ is the effect of genotype on disease risk. Given observed data $\{\boldsymbol{y}_i, \boldsymbol{g}_i\}$, estimates of the effect parameters, denoted $\{\hat{a}_i, \hat{b}_i\}$, are typically derived by maximizing the log-likelihood function

$$L(a_i, b_i) = \log P(\boldsymbol{y}_i \mid a_i, b_i, \boldsymbol{g}).$$

That is, values for $\{a_i, b_i\}$ which satisfy:

$$\frac{\partial L(a_i, b_i)}{\partial b_i} = \boldsymbol{g}^T (\boldsymbol{y} - \pi_{y_i|\boldsymbol{g}}) = 0.$$

(3)

Assuming that the effect of genotype on disease risk is small, in the sense that $|b_i g| < 1$, it follows that

$$\pi_{y_i|g} = P(Y_i = 1 \mid g) = \frac{e^{(a_i+b_i g)}}{1 + e^{(a_i+b_i g)}}$$

$$= \frac{e^{a_i}}{1 + e^{a_i}} + \left(\frac{e^{a_i}}{(1 + e^{a_i})^2}\right) b_i g + \mathcal{O}({b_i}^2)$$

$$= a_i^* + b_i^* g + \mathcal{O}(b_i{}^2)$$
$$= \boldsymbol{\beta}_i \widetilde{\boldsymbol{g}} + \mathcal{O}(b_i{}^2),$$

(4)

where we have made the transformation of variables:

$$\boldsymbol{\beta}_i = (a_i^*, b_i^*),$$
$$\widetilde{\boldsymbol{g}} = (1, g)^T.$$

Hence, when $|b_i g| < 1$ the conditional probability of disease is well approximated by the linear predictor $\boldsymbol{\beta}_i \widetilde{\boldsymbol{g}}$. As it will be important later, the relationship between parameters in the logistic and the linear predictor is

$$b_i = \frac{b_i^*}{a_i^*(1 - a_i^*)} \quad \text{and} \quad a_i = \log\left(\frac{a_i^*}{1 - a_i^*}\right).$$

(5)

Combining equations (3) and (4), it follows that the score of the logistic model satisfies

$$\frac{\partial L(\boldsymbol{\beta}_i)}{\partial \boldsymbol{\beta}_i} = \widetilde{\boldsymbol{g}}_i{}^T\left(\boldsymbol{y}_i - \pi_{y_i|\widetilde{\boldsymbol{g}}_i}\right) \approx \widetilde{\boldsymbol{g}}_i{}^T(\boldsymbol{y}_i - \boldsymbol{\beta}_i \widetilde{\boldsymbol{g}}_i), \qquad |b_i g_{ij}| < 1, \quad j = 1,2,\ldots,N_i,$$

which is approximately zero when

$$\widetilde{\boldsymbol{g}}_i{}^T(\boldsymbol{y}_i - \boldsymbol{\beta}_i \widetilde{\boldsymbol{g}}_i) = 0$$
$$\Rightarrow \quad \widehat{\boldsymbol{\beta}}_i = \left(\widetilde{\boldsymbol{g}}_i{}^T \widetilde{\boldsymbol{g}}_i\right)^{-1} \widetilde{\boldsymbol{g}}_i{}^T \boldsymbol{y}_i,$$

(6)

note that these are the familiar ordinary least-squares estimates of main effect parameters in a linearized model. When the vector of genotypes has been centered, i.e., $\overline{\boldsymbol{g}} = 0$, it is straightforward to show that:

$$\hat{a}_i^* = \frac{\sum_{j=1}^{N_i} y_{ij}}{N_i} = \frac{N_i^*}{N_i} \quad \text{and} \quad \hat{\sigma}_{a_i}^* = \frac{\sqrt{\sum_{j=1}^{N_i} \pi_{y_{ij}|g_{ij}}\left(1 - \pi_{y_{ij}|g_{ij}}\right)}}{N_i},$$

$$\hat{b}_i^* = \frac{\sum_{j=1}^{N_i} y_{ij} g_{ij}}{\sum_{j=1}^{N_i} g_{ij}^2} \quad \text{and} \quad \hat{\sigma}_{b_i}^* = \frac{\sqrt{\sum_{j=1}^{N_i} \pi_{y_{ij}|g_{ij}}\left(1 - \pi_{y_{ij}|g_{ij}}\right) g_{ij}^2}}{\sum_{j=1}^{N_i} g_{ij}^2}.$$

(7)

We have used $N_i^*$ to denote the number of cases in the $i$th study, thus $\hat{a}_i^*$ is a measure of population baseline disease prevalence. The linearized genetic effect in equation (4), however, can be interpreted as

$$\hat{b}_i^* = \frac{\sum_{j=1}^{N_i} y_{ij} g_j}{\sum_{j=1}^{N_i} g_j^2} = \frac{MAF_i^*}{MAF_i(1 - MAF_i)} \frac{N_i^*}{(N_i - 1)}, \qquad N_i \gg 1,$$

(8)

where $MAF_i^*$ is an estimate of the $MAF_i$ in the cases only:

$$MAF_i^* = \frac{1}{2N_i^*} \sum_{j=1}^{N_i} g_{ij} I(y_{ij} = 1).$$

(9)

We will use equations (8) and (9) later, when aiming to interpret uplift via core model parameters.

Recall the relationship between the parameters in the logistic and linearized models presented in equation (5). After an application of the delta method, the variance of $\hat{b}_i$ in the logistic model (1) can be written as:

$$\hat{\sigma}_{\hat{b}_i}^2 = \operatorname{var}\left(\frac{\hat{b}_i^*}{\hat{a}_i^*(1 - \hat{a}_i^*)}\right) = \left|\frac{\hat{\sigma}_{b_i}^*}{\hat{a}_i^*(1 - \hat{a}_i^*)}\right|^2 \left(1 + \mathcal{O}\left(\frac{\hat{b}_i^*}{\hat{a}_i^*(1 - \hat{a}_i^*)} \frac{\hat{\sigma}_{a_i}^*}{\hat{\sigma}_{b_i}^*}\right)\right).$$

(10)

On combining equations (5) and (7), we reveal that the Z-score of the genetic effect in the logistic model (1) is well approximated by the estimated Z-score from the linearized effects (equation (7)), i.e.,

$$Z_i = \frac{\hat{b}_i}{\hat{\sigma}_{\hat{b}_i}} \approx \frac{\frac{\hat{b}_i^*}{\hat{a}_i^*(1 - \hat{a}_i^*)}}{\left|\frac{\hat{\sigma}_{b_i}^*}{\hat{a}_i^*(1 - \hat{a}_i^*)}\right|} = \frac{\hat{b}_i^*}{\hat{\sigma}_{b_i}^*} \operatorname{sgn}\big(\hat{a}_i^*(1 - \hat{a}_i^*)\big) = Z_i^*,$$

where we have used the fact that $\operatorname{sgn}\big(a_i^*(1 - a_i^*)\big) = 1$ from equation (7). Thus,

$$Z_i \approx Z_i^*$$

(11)

and it follows therefore, that the uplift $\alpha$ can be written as:

$$\alpha = \frac{1 + \frac{\hat{\sigma}_1 Z_2}{\hat{\sigma}_2 Z_1}}{\sqrt{1 + \hat{\sigma}_1^2 / \hat{\sigma}_2^2}}$$

$$\approx \frac{1 + \frac{\hat{a}_2^*(1-\hat{a}_2^*)\hat{\sigma}_{b_1}^* Z_2^*}{\hat{a}_1^*(1-\hat{a}_1^*)\hat{\sigma}_{b_2}^* Z_1^*}}{\sqrt{1 + \left(\frac{\hat{a}_2^*(1-\hat{a}_2^*)\hat{\sigma}_{b_1}^*}{\hat{a}_1^*(1-\hat{a}_1^*)\hat{\sigma}_{b_2}^*}\right)^2}}.$$

(12)

**Explicit description of MAF enrichment on IVW uplift ($\alpha$)**

In this section our goal is to gain some intuition as to how uplift can vary in terms of core parameters, e.g., $MAF_i$, $N_i$, $N_i^*$, for each study $i \in \{1,2\}$. We have demonstrated that equation (12) provides an accurate approximation to uplift (see **Extended Data Figure 5, Supplementary Figure 2**), however these core parameters are implicitly defined in the approximation. To help provide some guidance on their explicit role, we replace $\hat{a}_i^*, \hat{\sigma}_{b_i}^*$ and $Z_i^*$ in equation (12) with large sample (population) estimates, i.e., on assuming $N_i \gg 1$ for $i \in \{1,2\}$. In addition, we make the assumption of weak genetic effects (i.e., $|b_i| < 1$), low disease prevalence ( $\pi_i \ll 1$) and rare/low MAF (in both studies), so that:

$$\hat{a}_i^*(1-\hat{a}_i^*) \approx \hat{a}_i^* \approx \pi_i \; : \quad N_i \gg 1 \text{ and } \pi_i \ll 1,$$

$$\pi_{y_{ij}|g_{ij}}\left(1-\pi_{y_{ij}|g_{ij}}\right) \approx \pi_{y_{ij}|g_{ij}} \; : \; \pi_{y_{ij}|g_{ij}} \ll 1,$$

$$\sum_{j=1}^{N_i} g_{ij}^2 \approx 2N_i MAF_i(1-MAF_i)$$

$$\approx 2N_i MAF_i, \quad N_i \gg 1.$$

(13)

Let $\widetilde{MAF}_i$ denote the MAF in the cases, i.e.,

$$\widetilde{MAF}_i = 0.5\sum_{j=1}^{N_i} I(y_{ij}=1)g_{ij},$$

then, in combination with equations (4) and (7), it follows that

$$\pi_{y_{ij}|g_{ij}} \approx \pi_i\left(1 + \frac{\widetilde{MAF}_i}{MAF_i}\right), \qquad N_i \gg 1 \text{ and } |b_i| < 1,$$

which, with equations (7), (12) and (13), returns:

$$\alpha \approx \frac{1 + \kappa \frac{\widetilde{MAF}_2}{\widetilde{MAF}_1}}{\sqrt{1 + \kappa \frac{MAF_2}{MAF_1}}}, \qquad N_i \gg 1 \text{ and } i \in \{1,2\},$$

(14)

where

$$\kappa = \frac{\pi_2}{\pi_1} \frac{N_2}{N_1} \left( \frac{1 + \frac{\widetilde{MAF}_1}{MAF_1}}{1 + \frac{\widetilde{MAF}_2}{MAF_2}} \right).$$

Note that $\tilde{X} = \frac{\widetilde{MAF}_2}{\widetilde{MAF}_1}$ denotes the $\tilde{X}$-fold enrichment of MAF in the cases and $X = \frac{MAF_2}{MAF_1}$ the $X$-fold enrichment in the base-line population. Equation (14) makes clear the connection between IVW uplift and MAF-enrichment in the additional study. While the general relationship between the core parameters is complex, equation (14) highlights broadly, that for fixed $\kappa$ IVW uplift $\alpha$ increases with increasing MAF-enrichment. A detail highlighted by both our theoretical and simulated results (**Extended Data Figure 5, Supplementary Figure 2**).

**MAF enrichment in the larger study:** $N_1 \geq N_2$ and $MAF_1 \geq MAF_2$.

From equation (14), if $MAF_1 \geq MAF_2$ and $\widetilde{MAF}_1 \geq \widetilde{MAF}_2$ then both $X \leq 1$ and $\tilde{X} \leq 1$ and thus uplift decreases as a function of increased MAF enrichment in study 1.

#### Supplementary Results: Simulations of MAF enrichment effect on inverse-variance weighted meta-analysis Z-scores

To go some way toward assessing the impact of MAF-enrichment for rare variants (MAF<1%) on IVW uplift in a realistic setting, we performed a simulation study informed by UKB and FG study information. We simulated two binary variables, representing two disease endpoints, with study sample sizes set to $N_1 = 392814$ and $N_2 = 260405$, respectively. To specify disease prevalence parameters, we computed the median ratio of disease prevalence across all $l = 1,2,\ldots,744$ disease endpoints in UKB and FG datasets, i.e., $\text{median}\left({}^{\pi_{2l}}/_{\pi_{1l}}\right)$ (**Supplementary Table 3**). As $\text{median}\left({}^{\pi_{2l}}/_{\pi_{1l}}\right) = 1.5$, we set $\pi_2 = 1.5\pi_1$ and fixed $\pi_1 = 0.005$, which is approximately 2 times the median disease prevalence across the 744 disease endpoints within UKB. To assess impact of MAF enrichment across a range of rare/low MAFs, we varied MAF in the reference study, $MAF_1 \in \{10^{-4}, 5 \times 10^{-4}, 10^{-3}, 2.5 \times 10^{-3}, 5 \times$

$10^{-3}, 0.01\}$ and X-fold MAF-enrichment in study 2, $X \in \{1, 5, 10, 20, 30, 50\}$. Note that $MAF_1 = 0.01$ and $X = 50$ results in $MAF_2 = 0.5$, at which point the effect allele switches in study 2. This motivated our choice of maximum $MAF$ and $X$-fold enrichment values.

Data for disease status were generated under a logistic model, i.e., equation (2):

$$logit\left(\pi_{y_i|g_{ij}}\right) = a_i + b_i * g_{ij},$$

$$a_i \approx logit(\pi_i),$$

where

$$\pi_{y_i|g_{ij}} = P(Y_i = 1 \mid g_{ij}), \qquad i \in \{1,2\}, j = 1,2, \dots, N_i,$$

denotes the probability of disease conditional on genotype $g_{ij}$ for the $j$th participant in the $i$th study. Finally, a value for $b_i$ - the effect of genotype on disease – was randomly sampled from the set of positive regression coefficients (to reflect the fact that vast majority of low frequency variants have disease risk-increasing effects), computed in UKB (for $b_1$ values) and FG (for $b_2$ values) (**Supplementary Table 3).**

We validate the accuracy of the theoretical approximation to the IVW uplift, i.e., equation (12), by monitoring the median absolute relative error (MARE) across $i = 1,2, \dots, 1000$ simulated datasets and over variety of MAF and enrichment values. Specifically, we computed

$$MARE_{jk} = \text{median}\left|\Omega_{jk}\right|, \quad \Omega_{jk} = \left\{\omega_{1jk}, \omega_{2jk}, \dots, \omega_{1000jk}\right\},$$

where

$$\omega_{ijk} = \left\{\frac{\alpha_{obs_{ijk}} - \alpha_{\exp_{ijk}}}{\alpha_{obs_{ijk}}}\right\}$$

with $j$ indexing the set of values $MAF_1 \in \{10^{-4}, 5 \times 10^{-4}, 10^{-3}, 5 \times 10^{-3}, 0.01\}$ and $k$ indexing the set of $X$-fold enrichment values $X \in \{1, 5, 10, 20, 30, 50\}$, i.e., $X = \frac{MAF_2}{MAF_1}$. Our findings reveal very good correspondence between the theoretical prediction (equation (12)) and simulated uplift $\alpha$-values (**Extended Data Figure 5 right**; **Supplementary Figure 2c**). Excluding the scenarios in which $MAF_1 = 10^{-4}$, the MARE (as a percentage) was $\lesssim 5$ % across the range of MAF and enrichment values considered: highlighting the accuracy of the approximation in equation (12) over a wide range or rare MAFs and MAF-enrichment values. The observed increase in MARE when $j = 10^{-4}$ is a consequence of the approximation equation (10) becoming less accurate – this happens when $N_1^* MAF_1 \lesssim 1$ and hence the number of cases with an effect allele in the reference sample is typically very small. **Extended Data**

**Figure 5** (and **Supplementary Figure 2)** makes clear that, on approaching and passing this boundary (which in our simulation set-up occurs when $MAF_1 \approx 5 \times 10^{-4}$) the MARE quickly increases. Despite this, in our simulation the MARE$\lesssim 20\%$ at $MAF_1 = 10^{-4}$.

#### Supplementary Results: Summary of theoretical and simulation results

We observed the uplift to increase with increasing MAF and allelic enrichment as expected. The gain in IVW uplift is not linear across MAFs, with most of the gain in uplift occurring at MAF<0.5% before plateauing (**Extended Data Figure 5, left; Supplementary Figure 2a**). There is a rapid increase in uplift as baseline MAF increases from $10^{-4}$ to $10^{-3}$ suggesting the added potential for novel findings benefitting very rare allele frequency ranges. The increase in utility at rare MAF ranges has practical impact on novel association findings as common and low frequency variant association can be well-powered to be detected already at biobank sizes for prevalent diseases. IVW uplift also increases with allelic enrichment, once again sharp increases in uplift are expected and observed at lower enrichment values, eventually flattening out towards MAF 1% and fold enrichment 50x (**Extended Data Figure 5, Supplementary Figure 2**). Note that, MAF ~1% and fold enrichment ~50x would fall within the region of inflection (top right quadrants of **Extended Data Figure 5a and 5b**) where the MAF in the enriched cohort approaches 50%. In practice, the vast majority of rare variants do not fall near the region of inflection as it would suggest, for example, a MAF of 1% in one study and near 50% in the other. Our results make clear that the surface of IVW uplift values using observed data is in general similar to theoretical results. Taken together, the simulation results both theoretically and empirically show the added utility in boosting association findings through cohort specific allelic enrichment additional to increased power from larger sample size.

#### Supplementary Methods: Functional characterization of PITX2c Pro41Ser

Methods relating to **Independent AF participants, DNA extraction, PCR and Sanger sequencing** relating to the analysis of association of Pro41Ser PITX2 with AF in a French cohort were previously described in[1].

##### *PITX2c* cDNA constructs

For gene reporter essay in TM-1 cells (Transformed human trabecular meshwork cells), the Pro41Ser variant was generated by site-directed mutagenesis (QuikChange®, Agilent Technologies, Mississauga, ON) using a pcDNA4/HisMax© plasmid (Invitrogen, Burlington, ON) expressing the wild-type Xpress™-tagged *PITX2c*[2]. The forward mutagenesis primers are similar to those reported before[1]. NdeI/XhoI restricted fragments were subcloned into pcDNA4:*PITX2c*. The plasmids were purified using QIAGEN Maxiprep or QIAprep® Spin Miniprep kits (QIAGEN Inc., Mississauga, ON) and validated by sequencing. For HL-1 cells experiments, we generated a V5-Tag-pcDNA3.1/Zeo vector carrying the open reading frames from human *PITX2* wild type and mutant sequences as previously described[1].

##### Western blot Analysis

Protein expression of the Pro41Ser variant was assessed in TM-1 cells using Anti-Xpress™ (Invitrogen) antibodies to detect Xpress-tagged recombinant proteins[3]. The PITX2c protein expression was also confirmed in HL-1 cells (mouse immortalized atrial myocardial cells)[4], transfected with V5-tagged vectors. The cells were seeded in 24-well plates and transfected using lipofectamine 2000 (Invitrogen) and 0.2µg of plasmid per well (V5-PITX2c WT or V5-PITX2c Pro41Ser). 24hr after transfection, cells were collected, and Western blot was performed as previously described[1].

##### Transactivation assays

PITX2c-expressing plasmid or the empty expression vector were co-transfected (n=3) with pGL3Basic-SLC13A3 reporter[2] and pCMVβ transfection control vector (Clontech Laboratories, Inc, Mountain View, CA) into TM-1 cells (500 ng, 60 ng and 30 ng respectively). Luciferase activity was measured by luminometry (Turner Designs, Sunnyvale, CA) according to the manufacturer's instructions and standardized to the β-galactosidase (internal control, β-galactosidase Enzyme Assay System, Promega, Madison, WI, USA).

**Immunolocalization**

Immunofluorescence was carried out as previously described[5]. 24hr after transfection, TM-1 cells (grown on coverslips) were fixed with 2% paraformaldehyde/PBS for 20 min. Antibodies: anti-Xpress and Mouse-Cy3 (Jackson ImmunoResearch, West Grove, PA) were used at a dilution of 1:500. The nuclei were labelled with a mounting medium containing DAPI. No significant difference of PITX2c protein expression was seen when normalized to GADPH internal control.

**Electrophoretic mobility shift assays**

WT and mutated PITX2c proteins' affinity with the DNA bicoid binding site were investigated by non-denaturing polyacrylamide gel electrophoresis of a mixture of double-stranded Cy3-labelled DNA (at 0.5 μM) and whole-cell extracts of TM-1 cells (40 μg) transfected with a *PITX2c* construct.

**Quantitative Reverse Transcriptase–PCR Analyses in HL-1 cells**

HL-1 cells transfected with V5-tagged vectors were collected 24h after transfection. Total RNA extraction was carried out using SV Total RNA isolation system (Promega) followed by cDNA synthesis using Maxima first strand cDNA synthesis kit for RT-qPCR (Fermentas life science) according to the manufacturers' instructions. qRT-PCR was performed using SsoFast™ EvaGreen (BioRad) supermix in a CFX384 QPCR System (BioRad).

All experimental quantitative data were analyzed using unpaired two-tailed Student's t-test. Unless otherwise mentioned, assays and measurements were done in triplicate. Data are reported as mean ± SEM.

#### Supplementary Results on PITX2c Pro41Ser

We genotyped the *PITX2* gene in a cohort of 60 unrelated patients with a history of atrial fibrillation and 389 controls. We identified a novel heterozygous variant, a single-base substitution of a cytosine by a thymidine at nucleotide 121 (c.121C>T) resulting in an amino acid change from proline to serine at position 41 (Pro41Ser) (**Supplementary figure Pro41Ser 1**). The propositus is a 65-years old female. She was first diagnosed with paroxysmal AF without heart disease when she was 48 years old.

##### Reporter gene assay

We verified that the increased transactivation effects did not result from a change in the intracellular distribution of the recombinant proteins since the nuclear localization was similar to that of the WT (**Supplementary figure Pro41Ser 2a**). Moreover, they were neither linked to a higher protein expression of the variant (not shown) nor to the ability of WT and mutated PITX2c proteins to interact with the DNA bicoid binding site as tested by EMSA assay (**Supplementary figure Pro41Ser 2b**). The full images of the EMSA gel and corresponding Western blot are shown in (**Supplementary figure Pro41Ser 3**).

##### qRT-PCR determination of connexins and ion channels

Control experiments show the differences in expression of PITX2c (**Supplementary figure Pro41Ser 4a**) that were taken into account to normalize the mRNA level measurements. Further, it was verified that cells transfected with an empty vector had mRNA levels similar to those in non-transfected cells (**Supplementary figure Pro41Ser 4b**).

#### Supplementary Discussion on PITX2c Pro41Ser

PITX2c Pro41Ser is the second PITX2c gain-of-function mutation identified as underlying AF and appears to impact PITX2c in a similar manner as a previously described Met207Val mutation that had been identified in a family study[1]. The Pro41Ser variant is situated in exon 4, which encodes a 52 amino-acid N-terminal sequence specific to PITX2c and humans. This domain has been ascribed a crucial role in cardiac asymmetric morphogenesis[6]. Its overexpression on the left side of the chick embryo randomized the direction of heart looping, an effect that was lost when a Lysine at amino acid position 41 (amino acid position 33 in human PITX2c) was artificially mutated to an Arginine[7].

We observed a gain-of-function in the transactivation activity of Pro41Ser-PITX2c. PITX2c has been found to be over-expressed in human atrial myocytes isolated from patients with chronic atrial fibrillation[8]. The Pro41Ser mutant in HL-1 cells increased KCNH2 mRNA 1.81-fold. Such an increase in the rapid outward rectifier current $I_{Kr}$ (encoded by KCNH2), likely shortens atrial action potential duration and refractory period, which favors re-entry of excitation and thus AF[9]. Such $I_{Kr}$ gain-of-function induced by mutations of KCNH2 has been associated with familial AF[10,11]. In HL-1 cells, the Pro41Ser mutant caused an increase in Cx40 and Cx43 mRNAs by1.76-fold and1.85-fold respectively. Such a parallel increase was found in Cx40 and Cx43 proteins in the left atrial myocardium of patients with AF[12]. In summary, the results obtained for Pro41Ser PITX2c experimentally are likely to explain the association of this variant with increased risk of AF.

### Supplementary Figures Pro41Ser PITX2c

**Supplementary Figure Pro41Ser 1. Electropherogram of the proband carrying the Pro41Ser variant.** Arrows show the cytosine to thymidine change and box shows the resulting codon change.

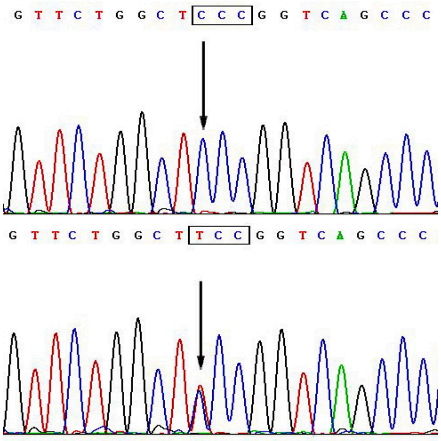

**Supplementary Figure Pro41Ser 2. (a) Immunofluorescence detection of recombinant PITX2c in TM-1 cells.** Merged images of DAPI-labelled nuclei (blue colour) and anti-PITX2c labelling with Anti-Xpress Ab (1:500) as detected by goat Anti-Mouse-IgG coupled to Cyanine3 (1:500) (red colour) in cells transfected with the WT cDNA (left) or the Pro41Ser variant cDNA (right). Red labelling was undetectable outside of nuclear location in TM-1 cells. The scale bar is 20μm**. (b) Upper panel: Electrophoretic mobility shift assays (EMSA) analysis.** Whole-cell extracts from TM-1 cells transfected with cDNA4: Xpress-PITX2c plasmids. Negative control: probe alone (**P**) or with empty vector (**P+EV**) exhibited a single low-mobility shifted background band. Complexes of probe bound to recombinant PITX2c constructs (**WT** and **Pro41Ser**), separated by blank lanes (**B**), were shifted to the same location in the respective lanes, with unbound probe migrating to the bottom of the gel. **Lower panel**: Western analysis shows the expression levels of the Xpress-PITX2c constructs, PITX2c WT (**WT**) and PITX2c Pro41Ser (**Pro41Ser**) in TM-1 cells, in relation to the α-tubulin internal lane control.

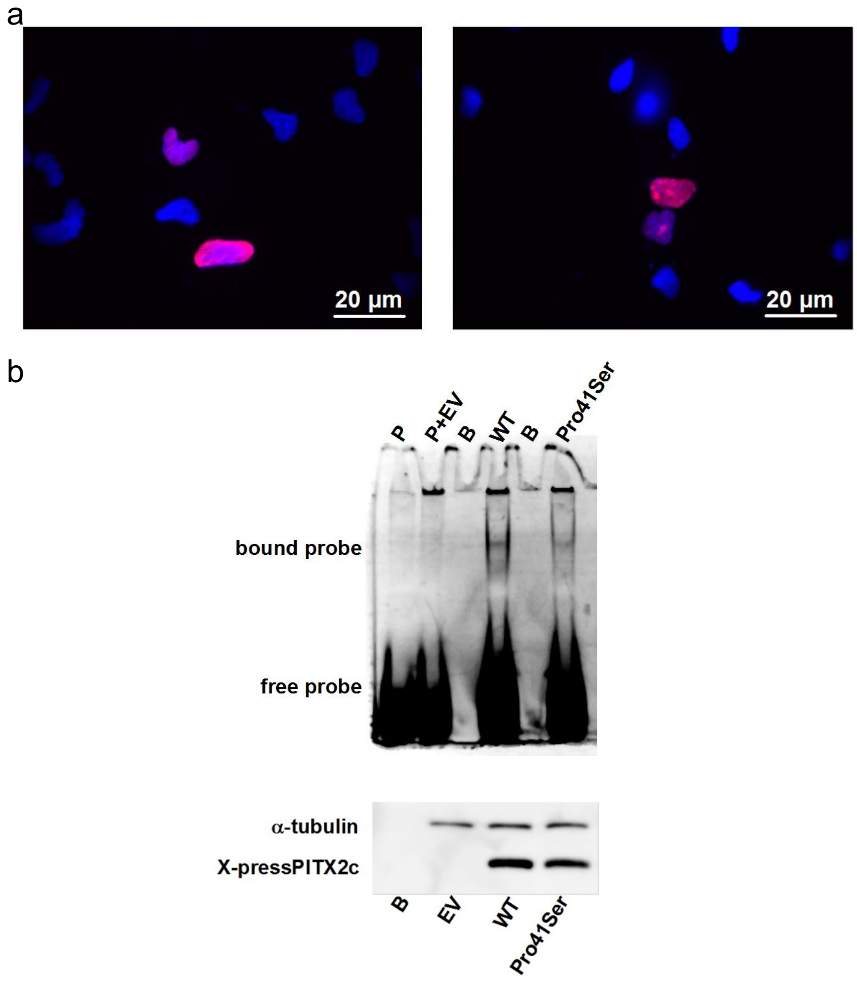

**Supplementary Figure Pro41Ser 3. Full size images of the EMSA gel.** (**a**) and corresponding western blot (**b**). **P**: probe only; **P+WT**: probe + PITX2c WT; **B**: blank; **WT**: PITX2c WT; **EV**: empty vector; **Pro41Ser**: PITX2c Pro41Ser; **NR**: not relevant to this study. This figure is provided for showing the absence of undue image manipulation for producing **Supplementary Figure Pro41Ser 2.**

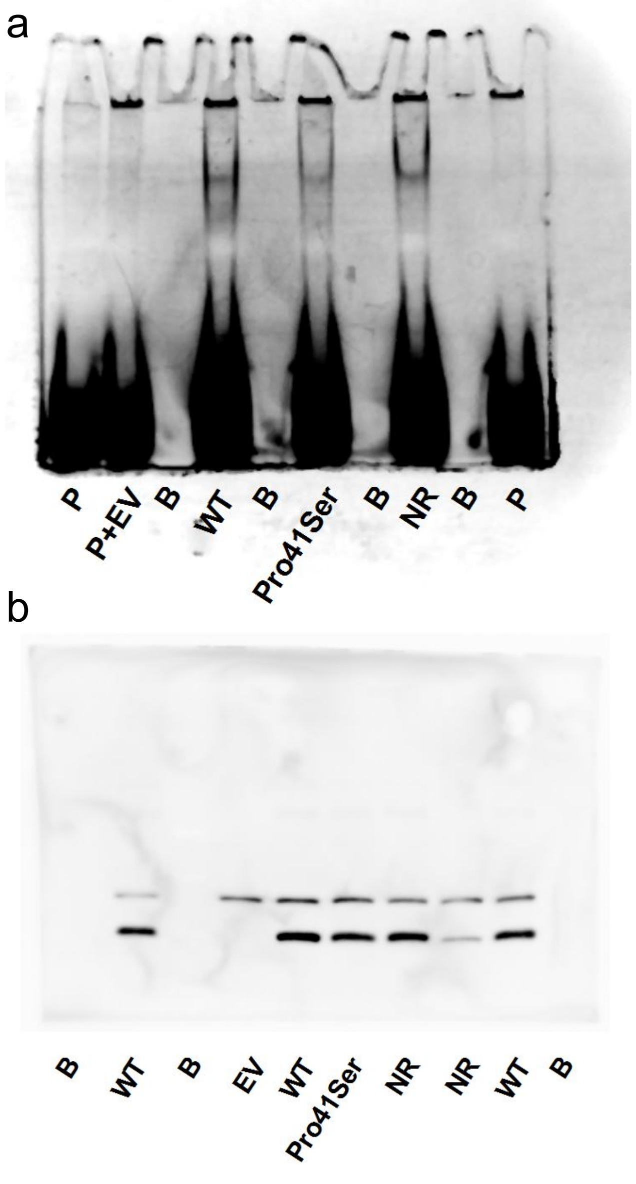

**Supplementary Figure Pro41Ser 4. (a) Quantification of PITX2c expression** in HL-1 cells transfected with either the **WT** or the Pro41Ser-mutated PITX2c plasmid (**Pro41Ser**) as normalized to GADPH internal control. The transfection experiments were repeated three times for semi-quantitative measurements of PITX2c expression. **(b) Levels of mRNA for connexins and ion channels in HL-1 cells**, either non-transfected (**NT**) or transfected with an empty vector (**EV**) (i.e: the V5-Tag-pcDNA3.1/Zeo vector devoid of any PITX2 sequence). Note that mRNA levels were not affected by transfection with the empty vector. Transfections and controls were repeated on three separate cell cultures. mRNA measurements were repeated three times on each mRNA extract.

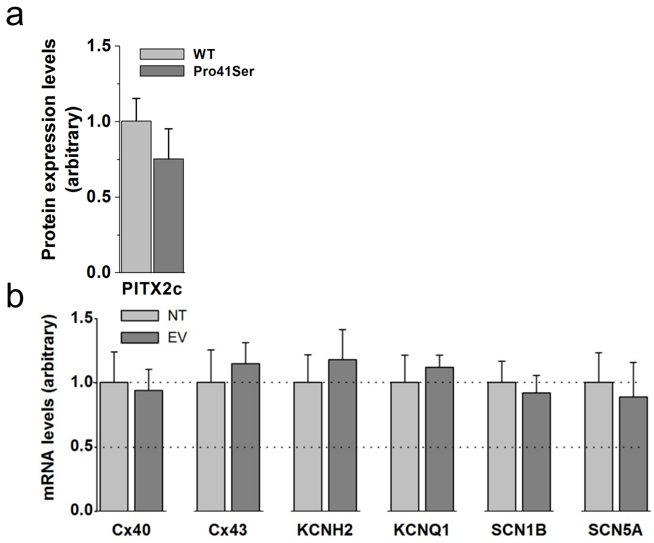

#### Supplementary Figure 1 Legend

**Interactive Manhattan plot summary of novel sentinel associations**. Size of the point is proportional to effect size. $-\log_{10}(p)$ capped at $-\log_{10}(10^{-50})$. Colours indicate disease groups (click [select/deselect trace]/double click [isolate one trace] on the legend to toggle selection). "+" indicate novel variant and gene, "×" indicate novel variant ($r^2<0.2$) not reported in GWAS Catalog/PhenoScanner for the disease. Dotted horizontal lines indicate $-\log_{10}(2x10^{-9})$ [brown] and $-\log_{10}(5x10^{-8})$ [grey]. Hover over the points for detailed information and double click/click on colors/shape in the legends to filter on select groups. Tooltip is available at top right for additional interactive options including: zooming and panning, selection, toggling multiple highlighting of nearby regions on hover and saving as static images.

#### Supplementary Figure 2 Legend

**Interactive surface plot of effects of cohort specific allele enrichment on inverse variant weighted meta-analysis z-scores (IVW uplift) across MAFs** (up to MAF 1%). Uplift is defined as the ratio of meta-analysed IVW Z-score to the Z-score of an individual study. **(a) theoretically predicted IVW uplift. (b) observed IVW uplift. (c) Median absolute relative error (MARE, %) between simulated and theoretical IVW uplift values.** For each combination of MAF and allelic enrichment, we simulated 1000 datasets for two binary variables reflecting disease status. Study sample size and disease prevalence were fixed (matching values estimates from UKB and FG), genomic effects were randomly sampled from the set of positive effect sizes in UKB and FG **(Supplementary Table 3)**, MAF was varied from 0.01% to 1% and allele enrichment (in the smaller study) ranged from 1 to 50. Tooltip is available at top right for additional interactive options including zooming, panning and saving as static images.

#### Biobank contributions to FinnGen

Auria Biobank (www.auria.fi/biopankki),

THL Biobank (www.thl.fi/biobank)

Helsinki Biobank (www.helsinginbiopankki.fi)

Biobank Borealis of Northern Finland (https://www.ppshp.fi/Tutkimus-ja-opetus/Biopankki/Pages/Biobank-Borealis-briefly-in-English.aspx)

Finnish Clinical Biobank Tampere (www.tays.fi/en-US/Research_and_development/Finnish_Clinical_Biobank_Tampere)

Biobank of Eastern Finland (www.ita-suomenbiopankki.fi/en)

Central Finland Biobank (www.ksshp.fi/fi-FI/Potilaalle/Biopankki)

Finnish Red Cross Blood Service Biobank (www.veripalvelu.fi/verenluovutus/biopankkitoiminta)

Terveystalo Biobank (www.terveystalo.com/fi/Yritystietoa/Terveystalo-Biopankki/Biopankki/)

All Finnish Biobanks are members of BBMRI.fi infrastructure (www.bbmri.fi) and FinBB (https://finbb.fi/).

#### FinnGen ethics statement details

Patients and control subjects in FinnGen provided informed consent for biobank research, based on the Finnish Biobank Act. Alternatively, separate research cohorts, collected prior the Finnish Biobank Act came into effect (in September 2013) and start of FinnGen (August 2017), were collected based on study-specific consents and later transferred to the Finnish biobanks after approval by Fimea, the National Supervisory Authority for Welfare and Health. Recruitment protocols followed the biobank protocols approved by Fimea. The Coordinating Ethics Committee of the Hospital District of Helsinki and Uusimaa (HUS) approved the FinnGen study protocol Nr HUS/990/2017.

The FinnGen study is approved by Finnish Institute for Health and Welfare (permit numbers: THL/2031/6.02.00/2017, THL/1101/5.05.00/2017, THL/341/6.02.00/2018, THL/2222/6.02.00/2018, THL/283/6.02.00/2019, THL/1721/5.05.00/2019, THL/1524/5.05.00/2020, and THL/2364/14.02/2020), Digital and population data service agency (permit numbers: VRK43431/2017-3, VRK/6909/2018-3, VRK/4415/2019-3), the Social Insurance Institution (permit numbers: KELA 58/522/2017, KELA 131/522/2018, KELA 70/522/2019, KELA 98/522/2019, KELA 138/522/2019, KELA 2/522/2020, KELA 16/522/2020 and Statistics Finland (permit numbers: TK-53-1041-17 and TK-53-90-20).

The Biobank Access Decisions for FinnGen samples and data utilized in FinnGen Data Freeze 6 include: THL Biobank BB2017_55, BB2017_111, BB2018_19, BB_2018_34, BB_2018_67, BB2018_71, BB2019_7, BB2019_8, BB2019_26, BB2020_1, Finnish Red Cross Blood Service Biobank 7.12.2017, Helsinki Biobank HUS/359/2017, Auria Biobank AB17-5154, Biobank Borealis of Northern Finland_2017_1013, Biobank of Eastern Finland 1186/2018, Finnish Clinical Biobank Tampere MH0004, Central Finland Biobank 1-2017, and Terveystalo Biobank STB 2018001.

### FinnGen consortium contributors

#### Steering Committee

Aarno Palotie Institute for Molecular Medicine Finland, HiLIFE, University of Helsinki, Finland
Mark Daly Institute for Molecular Medicine Finland, HiLIFE, University of Helsinki, Finland

**Pharmaceutical companies**

Bridget Riley-Gills Abbvie, Chicago, IL, United States
Howard Jacob Abbvie, Chicago, IL, United States
Dirk Paul Astra Zeneca, Cambridge, United Kingdom
Heiko Runz Biogen, Cambridge, MA, United States
Sally John Biogen, Cambridge, MA, United States
Robert Plenge Celgene, Summit, NJ, United States/Bristol Myers Squibb, New York, NY, United States
Mark McCarthy Genentech, San Francisco, CA, United States
Julie Hunkapiller Genentech, San Francisco, CA, United States
Meg Ehm GlaxoSmithKline, Brentford, United Kingdom
Kirsi Auro GlaxoSmithKline, Brentford, United Kingdom
Caroline Fox Merck, Kenilworth, NJ, United States
Anders Mälarstig Pfizer, New York, NY, United States
Katherine Klinger Sanofi, Paris, France
Deepak Raipal Sanofi, Paris, France
Tim Behrens Maze Therapeutics, San Francisco, CA, United States
Robert Yang Janssen Biotech, Beerse, Belgium
Richard Siegel Novartis, Basel, Switzerland

**University of Helsinki & Biobanks**

Tomi Mäkelä HiLIFE, University of Helsinki, Finland, Finland
Jaakko Kaprio Institute for Molecular Medicine Finland, HiLIFE, Helsinki, Finland, Finland
Petri Virolainen Auria Biobank / University of Turku / Hospital District of Southwest Finland, Turku, Finland
Antti Hakanen Auria Biobank / University of Turku / Hospital District of Southwest Finland, Turku, Finland
Terhi Kilpi THL Biobank / The National Institute of Health and Welfare Helsinki, Finland
Markus Perola THL Biobank / The National Institute of Health and Welfare Helsinki, Finland
Jukka Partanen Finnish Red Cross Blood Service / Finnish Hematology Registry and Clinical Biobank, Helsinki, Finland
Anne Pitkäranta Helsinki Biobank / Helsinki University and Hospital District of Helsinki and Uusimaa, Helsinki
Juhani Junttila Northern Finland Biobank Borealis / University of Oulu / Northern Ostrobothnia Hospital District, Oulu, Finland
Raisa Serpi Northern Finland Biobank Borealis / University of Oulu / Northern Ostrobothnia Hospital District, Oulu, Finland
Tarja Laitinen Finnish Clinical Biobank Tampere / University of Tampere / Pirkanmaa Hospital District, Tampere, Finland
Johanna Mäkelä Finnish Clinical Biobank Tampere / University of Tampere / Pirkanmaa Hospital District, Tampere, Finland
Veli-Matti Kosma Biobank of Eastern Finland / University of Eastern Finland / Northern Savo Hospital District, Kuopio, Finland
Urho Kujala Central Finland Biobank / University of Jyväskylä / Central Finland Health Care District, Jyväskylä, Finland

**Other Experts/ Non-Voting Members**

Outi Tuovila Business Finland, Helsinki, Finland
Raimo Pakkanen Business Finland, Helsinki, Finland

#### Scientific Committee

**Pharmaceutical companies**

Jeffrey Waring Abbvie, Chicago, IL, United States
Ali Abbasi Abbvie, Chicago, IL, United States
Mengzhen Liu Abbvie, Chicago, IL, United States
Ioanna Tachmazidou Astra Zeneca, Cambridge, United Kingdom
Chia-Yen Chen Biogen, Cambridge, MA, United States
Heiko Runz Biogen, Cambridge, MA, United States

Shameek Biswas Celgene, Summit, NJ, United States/Bristol Myers Squibb, New York, NY, United States
Julie Hunkapiller Genentech, San Francisco, CA, United States
Meg Ehm GlaxoSmithKline, Brentford, United Kingdom
Neha Raghavan Merck, Kenilworth, NJ, United States
Adriana Huertas-Vazquez Merck, Kenilworth, NJ, United States
Anders Mälarstig Pfizer, New York, NY, United States
Xinli Hu Pfizer, New York, NY, United States
Katherine Klinger Sanofi, Paris, France
Matthias Gossel Sanofi, Paris, France
Robert Graham Maze Therapeutics, San Francisco, CA, United States
Tim Behrens Maze Therapeutics, San Francisco, CA, United States
Beryl Cummings Maze Therapeutics, San Francisco, CA, United States
Wilco Fleuren Janssen Biotech, Beerse, Belgium
Dawn Waterworth Janssen Biotech, Beerse, Belgium
Nicole Renaud Novartis, Basel, Switzerland
Aviv Madar Novartis, Basel, Switzerland
Maen Obeidat Novartis, Basel, Switzerland

**University of Helsinki & Biobanks**

Samuli Ripatti Institute for Molecular Medicine Finland, HiLIFE, Helsinki, Finland
Johanna Schleutker Auria Biobank / Univ. of Turku / Hospital District of Southwest Finland, Turku, Finland
Markus Perola THL Biobank / The National Institute of Health and Welfare Helsinki, Finland
Mikko Arvas Finnish Red Cross Blood Service / Finnish Hematology Registry and Clinical Biobank, Helsinki, Finland
Olli Carpén Helsinki Biobank / Helsinki University and Hospital District of Helsinki and Uusimaa, Helsinki
Reetta Hinttala Northern Finland Biobank Borealis / University of Oulu / Northern Ostrobothnia Hospital District, Oulu, Finland
Johannes Kettunen Northern Finland Biobank Borealis / University of Oulu / Northern Ostrobothnia Hospital District, Oulu, Finland
Johanna Mäkelä Finnish Clinical Biobank Tampere / University of Tampere / Pirkanmaa Hospital District, Tampere, Finland
Arto Mannermaa Biobank of Eastern Finland / University of Eastern Finland / Northern Savo Hospital District, Kuopio, Finland
Jari Laukkanen Central Finland Biobank / University of Jyväskylä / Central Finland Health Care District, Jyväskylä, Finland
Urho Kujala Central Finland Biobank / University of Jyväskylä / Central Finland Health Care District, Jyväskylä, Finland

#### Clinical Groups

**Neurology Group**

Reetta Kälviäinen Northern Savo Hospital District, Kuopio, Finland
Valtteri Julkunen Northern Savo Hospital District, Kuopio, Finland
Hilkka Soininen Northern Savo Hospital District, Kuopio, Finland
Anne Remes Northern Ostrobothnia Hospital District, Oulu, Finland
Mikko Hiltunen Northern Savo Hospital District, Kuopio, Finland
Jukka Peltola Pirkanmaa Hospital District, Tampere, Finland
Pentti Tienari Hospital District of Helsinki and Uusimaa, Helsinki, Finland
Juha Rinne Hospital District of Southwest Finland, Turku, Finland
Roosa Kallionpää Hospital District of Southwest Finland, Turku, Finland
Ali Abbasi Abbvie, Chicago, IL, United States
Adam Ziemann Abbvie, Chicago, IL, United States
Jeffrey Waring Abbvie, Chicago, IL, United States
Sahar Esmaeeli Abbvie, Chicago, IL, United States
Nizar Smaoui Abbvie, Chicago, IL, United States
Anne Lehtonen Abbvie, Chicago, IL, United States
Susan Eaton Biogen, Cambridge, MA, United States
Heiko Runz Biogen, Cambridge, MA, United States
Sanni Lahdenperä Biogen, Cambridge, MA, United States

Janet van Adelsberg Celgene, Summit, NJ, United States/ Bristol Myers Squibb, New York, NY, United States
Shameek Biswas Celgene, Summit, NJ, United States/ Bristol Myers Squibb, New York, NY, United States
Julie Hunkapiller Genentech, San Francisco, CA, United States
Natalie Bowers Genentech, San Francisco, CA, United States
Edmond Teng Genentech, San Francisco, CA, United States
Sarah Pendergrass Genentech, San Francisco, CA, United States
Onuralp Soylemez Merck, Kenilworth, NJ, United States
Kari Linden Pfizer, New York, NY, United States
Fanli Xu GlaxoSmithKline, Brentford, United Kingdom
David Pulford GlaxoSmithKline, Brentford, United Kingdom
Kirsi Auro GlaxoSmithKline, Brentford, United Kingdom
Laura Addis GlaxoSmithKline, Brentford, United Kingdom
John Eicher GlaxoSmithKline, Brentford, United Kingdom
Minna Raivio Hospital District of Helsinki and Uusimaa, Helsinki, Finland
Sarah Pendergrass Genentech, San Francisco, CA, United States
Beryl Cummings Maze Therapeutics, San Francisco, CA, United States
Juulia Partanen Institute for Molecular Medicine Finland, HiLIFE, University of Helsinki, Finland

**Gastroenterology Group**

Martti Färkkilä Hospital District of Helsinki and Uusimaa, Helsinki, Finland
Jukka Koskela Hospital District of Helsinki and Uusimaa, Helsinki, Finland
Sampsa Pikkarainen Hospital District of Helsinki and Uusimaa, Helsinki, Finland
Airi Jussila Pirkanmaa Hospital District, Tampere, Finland
Katri Kaukinen Pirkanmaa Hospital District, Tampere, Finland
Timo Blomster Northern Ostrobothnia Hospital District, Oulu, Finland
Mikko Kiviniemi Northern Savo Hospital District, Kuopio, Finland
Markku Voutilainen Hospital District of Southwest Finland, Turku, Finland
Ali Abbasi Abbvie, Chicago, IL, United States
Graham Heap Abbvie, Chicago, IL, United States
Jeffrey Waring Abbvie, Chicago, IL, United States
Nizar Smaoui Abbvie, Chicago, IL, United States
Fedik Rahimov Abbvie, Chicago, IL, United States
Anne Lehtonen Abbvie, Chicago, IL, United States
Keith Usiskin Celgene, Summit, NJ, United States/ Bristol Myers Squibb, New York, NY, United States
Tim Lu Genentech, San Francisco, CA, United States
Natalie Bowers Genentech, San Francisco, CA, United States
Danny Oh Genentech, San Francisco, CA, United States
Sarah Pendergrass Genentech, San Francisco, CA, United States
Kirsi Kalpala Pfizer, New York, NY, United States
Melissa Miller Pfizer, New York, NY, United States
Xinli Hu Pfizer, New York, NY, United States
Linda McCarthy GlaxoSmithKline, Brentford, United Kingdom
Onuralp Soylemez Merck, Kenilworth, NJ, United States
Mark Daly Institute for Molecular Medicine Finland, HiLIFE, University of Helsinki, Finland

**Rheumatology Group**

Kari Eklund Hospital District of Helsinki and Uusimaa, Helsinki, Finland
Antti Palomäki Hospital District of Southwest Finland, Turku, Finland
Pia Isomäki Pirkanmaa Hospital District, Tampere, Finland
Laura Pirilä Hospital District of Southwest Finland, Turku, Finland
Oili Kaipiainen-Seppänen Northern Savo Hospital District, Kuopio, Finland
Johanna Huhtakangas Northern Ostrobothnia Hospital District, Oulu, Finland
Ali Abbasi Abbvie, Chicago, IL, United States
Jeffrey Waring Abbvie, Chicago, IL, United States
Fedik Rahimov Abbvie, Chicago, IL, United States
Apinya Lertratanakul Abbvie, Chicago, IL, United States
Nizar Smaoui Abbvie, Chicago, IL, United States
Anne Lehtonen Abbvie, Chicago, IL, United States
David Close Astra Zeneca, Cambridge, United Kingdom

Marla Hochfeld Celgene, Summit, NJ, United States/ Bristol Myers Squibb, New York, NY, United States
Natalie Bowers Genentech, San Francisco, CA, United States
Sarah Pendergrass Genentech, San Francisco, CA, United States
Onuralp Soylemez Merck, Kenilworth, NJ, United States
Kirsi Kalpala Pfizer, New York, NY, United States
Nan Bing Pfizer, New York, NY, United States
Xinli Hu Pfizer, New York, NY, United States
Jorge Esparza Gordillo GlaxoSmithKline, Brentford, United Kingdom
Kirsi Auro GlaxoSmithKline, Brentford, United Kingdom
Dawn Waterworth Janssen Biotech, Beerse, Belgium
Nina Mars Institute for Molecular Medicine Finland, HiLIFE, Helsinki, Finland

**Pulmonology Group**

Tarja Laitinen Pirkanmaa Hospital District, Tampere, Finland
Margit Pelkonen Northern Savo Hospital District, Kuopio, Finland
Paula Kauppi Hospital District of Helsinki and Uusimaa, Helsinki, Finland
Hannu Kankaanranta Pirkanmaa Hospital District, Tampere, Finland
Terttu Harju Northern Ostrobothnia Hospital District, Oulu, Finland
Riitta Lahesmaa Hospital District of Southwest Finland, Turku, Finland
Nizar Smaoui Abbvie, Chicago, IL, United States
Alex Mackay Astra Zeneca, Cambridge, United Kingdom
Glenda Lassi Astra Zeneca, Cambridge, United Kingdom
Susan Eaton Biogen, Cambridge, MA, United States
Steven Greenberg Celgene, Summit, NJ, United States/ Bristol Myers Squibb, New York, NY, United States
Hubert Chen Genentech, San Francisco, CA, United States
Sarah Pendergrass Genentech, San Francisco, CA, United States
Natalie Bowers Genentech, San Francisco, CA, United States
Joanna Betts GlaxoSmithKline, Brentford, United Kingdom
Soumitra Ghosh GlaxoSmithKline, Brentford, United Kingdom
Kirsi Auro GlaxoSmithKline, Brentford, United Kingdom
Rajashree Mishra GlaxoSmithKline, Brentford, United Kingdom
Sina Rüeger Institute for Molecular Medicine Finland, HiLIFE, University of Helsinki, Finland

**Cardiometabolic Diseases Group**

Teemu Niiranen The National Institute of Health and Welfare Helsinki, Finland
Felix Vaura The National Institute of Health and Welfare Helsinki, Finland
Veikko Salomaa The National Institute of Health and Welfare Helsinki, Finland
Markus Juonala Hospital District of Southwest Finland, Turku, Finland
Kaj Metsärinne Hospital District of Southwest Finland, Turku, Finland
Mika Kähönen Pirkanmaa Hospital District, Tampere, Finland
Juhani Junttila Northern Ostrobothnia Hospital District, Oulu, Finland
Markku Laakso Northern Savo Hospital District, Kuopio, Finland
Jussi Pihlajamäki Northern Savo Hospital District, Kuopio, Finland
Daniel Gordin Hospital District of Helsinki and Uusimaa, Helsinki, Finland
Juha Sinisalo Hospital District of Helsinki and Uusimaa, Helsinki, Finland
Marja-Riitta Taskinen Hospital District of Helsinki and Uusimaa, Helsinki, Finland
Tiinamaija Tuomi Hospital District of Helsinki and Uusimaa, Helsinki, Finland
Jari Laukkanen Central Finland Health Care District, Jyväskylä, Finland
Benjamin Challis Astra Zeneca, Cambridge, United Kingdom
Dirk Paul Astra Zeneca, Cambridge, United Kingdom
Julie Hunkapiller Genentech, San Francisco, CA, United States
Natalie Bowers Genentech, San Francisco, CA, United States
Sarah Pendergrass Genentech, San Francisco, CA, United States
Onuralp Soylemez Merck, Kenilworth, NJ, United States
Jaakko Parkkinen Pfizer, New York, NY, United States
Melissa Miller Pfizer, New York, NY, United States
Russell Miller Pfizer, New York, NY, United States
Audrey Chu GlaxoSmithKline, Brentford, United Kingdom
Kirsi Auro GlaxoSmithKline, Brentford, United Kingdom

Keith Usiskin Celgene, Summit, NJ, United States/ Bristol Myers Squibb, New York, NY, United States
Amanda Elliott Institute for Molecular Medicine Finland, HiLIFE, University of Helsinki, Finland / Broad Institute, Cambridge, MA, United States
Joel Rämö Institute for Molecular Medicine Finland, HiLIFE, University of Helsinki, Finland
Samuli Ripatti Institute for Molecular Medicine Finland, HiLIFE, University of Helsinki, Finland
Mary Pat Reeve Institute for Molecular Medicine Finland, HiLIFE, University of Helsinki, Finland
Sanni Ruotsalainen Institute for Molecular Medicine Finland, HiLIFE, University of Helsinki, Finland

**Oncology Group**

Tuomo Meretoja Hospital District of Helsinki and Uusimaa, Helsinki, Finland
Heikki Joensuu Hospital District of Helsinki and Uusimaa, Helsinki, Finland
Olli Carpén Hospital District of Helsinki and Uusimaa, Helsinki, Finland
Lauri Aaltonen Hospital District of Helsinki and Uusimaa, Helsinki, Finland
Johanna Mattson Hospital District of Helsinki and Uusimaa, Helsinki, Finland
Annika Auranen Pirkanmaa Hospital District , Tampere, Finland
Peeter Karihtala Northern Ostrobothnia Hospital District, Oulu, Finland
Saila Kauppila Northern Ostrobothnia Hospital District, Oulu, Finland
Päivi Auvinen Northern Savo Hospital District, Kuopio, Finland
Klaus Elenius Hospital District of Southwest Finland, Turku, Finland
Johanna Schleutker Hospital District of Southwest Finland, Turku, Finland
Relja Popovic Abbvie, Chicago, IL, United States
Jeffrey Waring Abbvie, Chicago, IL, United States
Bridget Riley-Gillis Abbvie, Chicago, IL, United States
Anne Lehtonen Abbvie, Chicago, IL, United States
Jennifer Schutzman Genentech, San Francisco, CA, United States
Julie Hunkapiller Genentech, San Francisco, CA, United States
Natalie Bowers Genentech, San Francisco, CA, United States
Sarah Pendergrass Genentech, San Francisco, CA, United States
Andrey Loboda Merck, Kenilworth, NJ, United States
Aparna Chhibber Merck, Kenilworth, NJ, United States
Heli Lehtonen Pfizer, New York, NY, United States
Stefan McDonough Pfizer, New York, NY, United States
Marika Crohns Sanofi, Paris, France
Sauli Vuoti Sanofi, Paris, France
Diptee Kulkarni GlaxoSmithKline, Brentford, United Kingdom
Kirsi Auro GlaxoSmithKline, Brentford, United Kingdom
Esa Pitkänen Institute for Molecular Medicine Finland, HiLIFE, University of Helsinki, Finland
Nina Mars Institute for Molecular Medicine Finland, HiLIFE, University of Helsinki, Finland
Mark Daly Institute for Molecular Medicine Finland, HiLIFE, University of Helsinki, Finland

**Ophthalmology Group**

Kai Kaarniranta Northern Savo Hospital District, Kuopio, Finland
Joni A Turunen Hospital District of Helsinki and Uusimaa, Helsinki, Finland
Terhi Ollila Hospital District of Helsinki and Uusimaa, Helsinki, Finland
Sanna Seitsonen Hospital District of Helsinki and Uusimaa, Helsinki, Finland
Hannu Uusitalo Pirkanmaa Hospital District, Tampere, Finland
Vesa Aaltonen Hospital District of Southwest Finland, Turku, Finland
Hannele Uusitalo-Järvinen Pirkanmaa Hospital District, Tampere, Finland
Marja Luodonpää Northern Ostrobothnia Hospital District, Oulu, Finland
Nina Hautala Northern Ostrobothnia Hospital District, Oulu, Finland
Mengzhen Liu Abbvie, Chicago, IL, United States
Heiko Runz Biogen, Cambridge, MA, United States
Stephanie Loomis Biogen, Cambridge, MA, United States
Erich Strauss Genentech, San Francisco, CA, United States
Natalie Bowers Genentech, San Francisco, CA, United States
Hao Chen Genentech, San Francisco, CA, United States
Sarah Pendergrass Genentech, San Francisco, CA, United States
Anna Podgornaia Merck, Kenilworth, NJ, United States
Juha Karjalainen Institute for Molecular Medicine Finland, HiLIFE, University of Helsinki, Finland / Broad Institute, Cambridge, MA, United States
Esa Pitkänen Institute for Molecular Medicine Finland, HiLIFE, University of Helsinki, Finland

**Dermatology Group**

| | |
|---|---|
| Kaisa Tasanen | Northern Ostrobothnia Hospital District, Oulu, Finland |
| Laura Huilaja | Northern Ostrobothnia Hospital District, Oulu, Finland |
| Katariina Hannula-Jouppi | Hospital District of Helsinki and Uusimaa, Helsinki, Finland |
| Teea Salmi | Pirkanmaa Hospital District, Tampere, Finland |
| Sirkku Peltonen | Hospital District of Southwest Finland, Turku, Finland |
| Leena Koulu | Hospital District of Southwest Finland, Turku, Finland |
| Kirsi Kalpala | Pfizer, New York, NY, United States |
| Ying Wu | Pfizer, New York, NY, United States |
| David Choy | Genentech, San Francisco, CA, United States |
| Sarah Pendergrass | Genentech, San Francisco, CA, United States |
| Nizar Smaoui | Abbvie, Chicago, IL, United States |
| Fedik Rahimov | Abbvie, Chicago, IL, United States |
| Anne Lehtonen | Abbvie, Chicago, IL, United States |
| Dawn Waterworth | Janssen Biotech, Beerse, Belgium |

**Odontology Group**

| | |
|---|---|
| Pirkko Pussinen | Hospital District of Helsinki and Uusimaa, Helsinki, Finland |
| Aino Salminen | Hospital District of Helsinki and Uusimaa, Helsinki, Finland |
| Tuula Salo | Hospital District of Helsinki and Uusimaa, Helsinki, Finland |
| David Rice | Hospital District of Helsinki and Uusimaa, Helsinki, Finland |
| Pekka Nieminen | Hospital District of Helsinki and Uusimaa, Helsinki, Finland |
| Ulla Palotie | Hospital District of Helsinki and Uusimaa, Helsinki, Finland |
| Juha Sinisalo | Hospital District of Helsinki and Uusimaa, Helsinki, Finland |
| Maria Siponen | Northern Savo Hospital District, Kuopio, Finland |
| Liisa Suominen | Northern Savo Hospital District, Kuopio, Finland |
| Päivi Mäntylä | Northern Savo Hospital District, Kuopio, Finland |
| Ulvi Gursoy | Hospital District of Southwest Finland, Turku, Finland |
| Vuokko Anttonen | Northern Ostrobothnia Hospital District, Oulu, Finland |
| Kirsi Sipilä | Northern Ostrobothnia Hospital District, Oulu, Finland |
| Sarah Pendergrass | Genentech, San Francisco, CA, United States |

**Women's Health and Reproduction Group**

| | |
|---|---|
| Hannele Laivuori | Institute for Molecular Medicine Finland, HiLIFE, University of Helsinki, Finland |
| Venla Kurra | Pirkanmaa Hospital District, Tampere, Finland |
| Oskari Heikinheimo | Hospital District of Helsinki and Uusimaa, Helsinki, Finland |
| Ilkka Kalliala | Hospital District of Helsinki and Uusimaa, Helsinki, Finland |
| Laura Kotaniemi-Talonen | Pirkanmaa Hospital District, Tampere, Finland |
| Kari Nieminen | Pirkanmaa Hospital District, Tampere, Finland |
| Päivi Polo | Hospital District of Southwest Finland, Turku, Finland |
| Kaarin Mäkikallio | Hospital District of Southwest Finland, Turku, Finland |
| Eeva Ekholm | Hospital District of Southwest Finland, Turku, Finland |
| Marja Vääräsmäki | Northern Ostrobothnia Hospital District, Oulu, Finland |
| Outi Uimari | Northern Ostrobothnia Hospital District, Oulu, Finland |
| Laure Morin-Papunen | Northern Ostrobothnia Hospital District, Oulu, Finland |
| Marjo Tuppurainen | Northern Savo Hospital District, Kuopio, Finland |
| Katja Kivinen | Institute for Molecular Medicine Finland, HiLIFE, University of Helsinki, Finland |
| Elisabeth Widen | Institute for Molecular Medicine Finland, HiLIFE, University of Helsinki, Finland |
| Taru Tukiainen | Institute for Molecular Medicine Finland, HiLIFE, University of Helsinki, Finland |
| Mary Pat Reeve | Institute for Molecular Medicine Finland, HiLIFE, University of Helsinki, Finland |
| Mark Daly | Institute for Molecular Medicine Finland, HiLIFE, University of Helsinki, Finland |
| Liu Aoxing | Institute for Molecular Medicine Finland, HiLIFE, University of Helsinki, Finland |
| Eija Laakkonen | University of Jyväskylä, Jyväskylä, Finland |
| Niko Välimäki | University of Helsinki, Helsinki, Finland |
| Lauri Aaltonen | Hospital District of Helsinki and Uusimaa, Helsinki, Finland |
| Johannes Kettunen | Northern Ostrobothnia Hospital District, Oulu, Finland |
| Mikko Arvas | Finnish Red Cross Blood Service, Helsinki, Finland |
| Jeffrey Waring | Abbvie, Chicago, IL, United States |
| Bridget Riley-Gillis | Abbvie, Chicago, IL, United States |
| Mengzhen Liu | Abbvie, Chicago, IL, United States |
| Janet Kumar | GlaxoSmithKline, Brentford, United Kingdom |
| Kirsi Auro | GlaxoSmithKline, Brentford, United Kingdom |

| | |
|---|---|
| Andrea Ganna | Institute for Molecular Medicine Finland, HiLIFE, University of Helsinki, Finland |
| Sarah Pendergrass | Genentech, San Francisco, CA, United States |

**FinnGen Analysis working group**

| | |
|---|---|
| Justin Wade Davis | Abbvie, Chicago, IL, United States |
| Bridget Riley-Gillis | Abbvie, Chicago, IL, United States |
| Danjuma Quarless | Abbvie, Chicago, IL, United States |
| Fedik Rahimov | Abbvie, Chicago, IL, United States |
| Sahar Esmaeeli | Abbvie, Chicago, IL, United States |
| Slavé Petrovski | Astra Zeneca, Cambridge, United Kingdom |
| Eleonor Wigmore | Astra Zeneca, Cambridge, United Kingdom |
| Adele Mitchell | Biogen, Cambridge, MA, United States |
| Benjamin Sun | Biogen, Cambridge, MA, United States |
| Ellen Tsai | Biogen, Cambridge, MA, United States |
| Denis Baird | Biogen, Cambridge, MA, United States |
| Paola Bronson | Biogen, Cambridge, MA, United States |
| Ruoyu Tian | Biogen, Cambridge, MA, United States |
| Stephanie Loomis | Biogen, Cambridge, MA, United States |
| Yunfeng Huang | Biogen, Cambridge, MA, United States |
| Joseph Maranville | Celgene, Summit, NJ, United States/ Bristol Myers Squibb, New York, NY, United States |
| Shameek Biswas | Celgene, Summit, NJ, United States/ Bristol Myers Squibb, New York, NY, United States |
| Elmutaz Mohammed | Celgene, Summit, NJ, United States/ Bristol Myers Squibb, New York, NY, United States |
| Samir Wadhawan | Celgene, Summit, NJ, United States/ Bristol Myers Squibb, New York, NY, United States |
| Erika Kvikstad | Celgene, Summit, NJ, United States/ Bristol Myers Squibb, New York, NY, United States |
| Minal Caliskan | Celgene, Summit, NJ, United States/ Bristol Myers Squibb, New York, NY, United States |
| Diana Chang | Genentech, San Francisco, CA, United States |
| Julie Hunkapiller | Genentech, San Francisco, CA, United States |
| Tushar Bhangale | Genentech, San Francisco, CA, United States |
| Natalie Bowers | Genentech, San Francisco, CA, United States |
| Sarah Pendergrass | Genentech, San Francisco, CA, United States |
| Kirill Shkura | Merck, Kenilworth, NJ, United States |
| Victor Neduva | Merck, Kenilworth, NJ, United States |
| Xing Chen | Pfizer, New York, NY, United States |
| Åsa Hedman | Pfizer, New York, NY, United States |
| Karen S King | GlaxoSmithKline, Brentford, United Kingdom |
| Padhraig Gormley | GlaxoSmithKline, Brentford, United Kingdom |
| Jimmy Liu | GlaxoSmithKline, Brentford, United Kingdom |
| Clarence Wang | Sanofi, Paris, France |
| Ethan Xu | Sanofi, Paris, France |
| Franck Auge | Sanofi, Paris, France |
| Clement Chatelain | Sanofi, Paris, France |
| Deepak Rajpal | Sanofi, Paris, France |
| Dongyu Liu | Sanofi, Paris, France |
| Katherine Call | Sanofi, Paris, France |
| Tai-He Xia | Sanofi, Paris, France |
| Beryl Cummings | Maze Therapeutics, San Francisco, CA, United States |
| Matt Brauer | Maze Therapeutics, San Francisco, CA, United States |
| Huilei Xu | Novartis, Basel, Switzerland |
| Amy Cole | Novartis, Basel, Switzerland |
| Jonathan Chung | Novartis, Basel, Switzerland |
| Jaison Jacob | Novartis, Basel, Switzerland |
| Katrina de Lange | Novartis, Basel, Switzerland |
| Jonas Zierer | Novartis, Basel, Switzerland |

Mitja Kurki Institute for Molecular Medicine Finland, HiLIFE, University of Helsinki, Finland / Broad Institute, Cambridge, MA, United States
Samuli Ripatti Institute for Molecular Medicine Finland, HiLIFE, University of Helsinki, Finland
Mark Daly Institute for Molecular Medicine Finland, HiLIFE, University of Helsinki, Finland
Juha Karjalainen Institute for Molecular Medicine Finland, HiLIFE, University of Helsinki, Finland / Broad Institute, Cambridge, MA, United States
Aki Havulinna Institute for Molecular Medicine Finland, HiLIFE, University of Helsinki, Finland
Juha Mehtonen Institute for Molecular Medicine Finland, HiLIFE, University of Helsinki, Finland
Priit Palta Institute for Molecular Medicine Finland, HiLIFE, University of Helsinki, Finland
Shabbeer Hassan Institute for Molecular Medicine Finland, HiLIFE, University of Helsinki, Finland
Pietro Della Briotta Parolo Institute for Molecular Medicine Finland, HiLIFE, University of Helsinki, Finland
Wei Zhou Broad Institute, Cambridge, MA, United States
Mutaamba Maasha Broad Institute, Cambridge, MA, United States
Shabbeer Hassan Institute for Molecular Medicine Finland, HiLIFE, University of Helsinki, Finland
Susanna Lemmelä Institute for Molecular Medicine Finland, HiLIFE, University of Helsinki, Finland
Manuel Rivas University of Stanford, Stanford, CA, United States
Aarno Palotie Institute for Molecular Medicine Finland, HiLIFE, University of Helsinki, Finland
Arto Lehisto Institute for Molecular Medicine Finland, HiLIFE, University of Helsinki, Finland
Andrea Ganna Institute for Molecular Medicine Finland, HiLIFE, University of Helsinki, Finland
Vincent Llorens Institute for Molecular Medicine Finland, HiLIFE, University of Helsinki, Finland
Hannele Laivuori Institute for Molecular Medicine Finland, HiLIFE, University of Helsinki, Finland
Mari E Niemi Institute for Molecular Medicine Finland, HiLIFE, University of Helsinki, Finland
Taru Tukiainen Institute for Molecular Medicine Finland, HiLIFE, University of Helsinki, Finland
Mary Pat Reeve Institute for Molecular Medicine Finland, HiLIFE, University of Helsinki, Finland
Henrike Heyne Institute for Molecular Medicine Finland, HiLIFE, University of Helsinki, Finland
Nina Mars Institute for Molecular Medicine Finland, HiLIFE, University of Helsinki, Finland
Kimmo Palin University of Helsinki, Helsinki, Finland
Javier Garcia-Tabuenca University of Tampere, Tampere, Finland
Harri Siirtola University of Tampere, Tampere, Finland
Tuomo Kiiskinen Institute for Molecular Medicine Finland, HiLIFE, University of Helsinki, Finland
Jiwoo Lee Institute for Molecular Medicine Finland, HiLIFE, University of Helsinki, Finland / Broad Institute, Cambridge, MA, United States
Kristin Tsuo Institute for Molecular Medicine Finland, HiLIFE, University of Helsinki, Finland / Broad Institute, Cambridge, MA, United States
Amanda Elliott Institute for Molecular Medicine Finland, HiLIFE, University of Helsinki, Finland / Broad Institute, Cambridge, MA, United States
Kati Kristiansson THL Biobank / The National Institute of Health and Welfare Helsinki, Finland
Mikko Arvas Finnish Red Cross Blood Service / Finnish Hematology Registry and Clinical Biobank, Helsinki, Finland
Kati Hyvärinen Finnish Red Cross Blood Service, Helsinki, Finland
Jarmo Ritari Finnish Red Cross Blood Service, Helsinki, Finland
Miika Koskinen Helsinki Biobank / Helsinki University and Hospital District of Helsinki and Uusimaa, Helsinki
Olli Carpén Helsinki Biobank / Helsinki University and Hospital District of Helsinki and Uusimaa, Helsinki
Johannes Kettunen Northern Finland Biobank Borealis / University of Oulu / Northern Ostrobothnia Hospital District, Oulu, Finland
Katri Pylkäs University of Oulu, Oulu, Finland
Marita Kalaoja University of Oulu, Oulu, Finland
Minna Karjalainen University of Oulu, Oulu, Finland
Tuomo Mantere Northern Finland Biobank Borealis / University of Oulu / Northern Ostrobothnia Hospital District, Oulu, Finland
Eeva Kangasniemi Finnish Clinical Biobank Tampere / University of Tampere / Pirkanmaa Hospital District, Tampere, Finland
Sami Heikkinen University of Eastern Finland, Kuopio, Finland
Arto Mannermaa Biobank of Eastern Finland / University of Eastern Finland / Northern Savo Hospital District, Kuopio, Finland
Eija Laakkonen University of Jyväskylä, Jyväskylä, Finland
Samuel Heron University of Turku, Turku, Finland
Dhanaprakash Jambulingam University of Turku, Turku, Finland

Venkat Subramaniam Rathinakannan University of Turku, Turku, Finland
Nina Pitkänen Auria Biobank / University of Turku / Hospital District of Southwest Finland, Turku, Finland

#### Biobank directors

Lila Kallio Auria Biobank / University of Turku / Hospital District of Southwest Finland, Turku, Finland
Sirpa Soini THL Biobank / The National Institute of Health and Welfare Helsinki, Finland
Jukka Partanen Finnish Red Cross Blood Service / Finnish Hematology Registry and Clinical Biobank, Helsinki, Finland
Eero Punkka Helsinki Biobank / Helsinki University and Hospital District of Helsinki and Uusimaa, Helsinki
Raisa Serpi Northern Finland Biobank Borealis / University of Oulu / Northern Ostrobothnia Hospital District, Oulu, Finland
Johanna Mäkelä Finnish Clinical Biobank Tampere / University of Tampere / Pirkanmaa Hospital District, Tampere, Finland
Veli-Matti Kosma Biobank of Eastern Finland / University of Eastern Finland / Northern Savo Hospital District, Kuopio, Finland
Teijo Kuopio Central Finland Biobank / University of Jyväskylä / Central Finland Health Care District, Jyväskylä, Finland

#### FinnGen Teams

**Administration**
Anu Jalanko Institute for Molecular Medicine Finland, HiLIFE, University of Helsinki, Finland
Huei-Yi Shen Institute for Molecular Medicine Finland, HiLIFE, University of Helsinki, Finland
Risto Kajanne Institute for Molecular Medicine Finland, HiLIFE, University of Helsinki, Finland
Mervi Aavikko Institute for Molecular Medicine Finland, HiLIFE, University of Helsinki, Finland
**Analysis**
Mitja Kurki Institute for Molecular Medicine Finland, HiLIFE, University of Helsinki, Finland / Broad Institute, Cambridge, MA, United States
Juha Karjalainen Institute for Molecular Medicine Finland, HiLIFE, University of Helsinki, Finland / Broad Institute, Cambridge, MA, United States
Pietro Della Briotta Parolo Institute for Molecular Medicine Finland, HiLIFE, University of Helsinki, Finland
Arto Lehisto Institute for Molecular Medicine Finland, HiLIFE, University of Helsinki, Finland
Juha Mehtonen Institute for Molecular Medicine Finland, HiLIFE, University of Helsinki, Finland
Wei Zhou Broad Institute, Cambridge, MA, United States
Masahiro Kanai Broad Institute, Cambridge, MA, United States
Mutaamba Maasha Broad Institute, Cambridge, MA, United States
**Clinical Endpoint Development**
Hannele Laivuori Institute for Molecular Medicine Finland, HiLIFE, University of Helsinki, Finland
Aki Havulinna Institute for Molecular Medicine Finland, HiLIFE, University of Helsinki, Finland
Susanna Lemmelä Institute for Molecular Medicine Finland, HiLIFE, University of Helsinki, Finland
Tuomo Kiiskinen Institute for Molecular Medicine Finland, HiLIFE, University of Helsinki, Finland
L. Elisa Lahtela Institute for Molecular Medicine Finland, HiLIFE, University of Helsinki, Finland
Matti Peura Institute for Molecular Medicine Finland, HiLIFE, University of Helsinki, Finland
**Communication**
Mari Kaunisto Institute for Molecular Medicine Finland, HiLIFE, University of Helsinki, Finland
**Data Management and IT Infrastructure**
Elina Kilpeläinen Institute for Molecular Medicine Finland, HiLIFE, University of Helsinki, Finland
Timo P. Sipilä Institute for Molecular Medicine Finland, HiLIFE, University of Helsinki, Finland
Georg Brein Institute for Molecular Medicine Finland, HiLIFE, University of Helsinki, Finland
Oluwaseun A. Dada Institute for Molecular Medicine Finland, HiLIFE, University of Helsinki, Finland
Awaisa Ghazal Institute for Molecular Medicine Finland, HiLIFE, University of Helsinki, Finland
Anastasia Shcherban Institute for Molecular Medicine Finland, HiLIFE, University of Helsinki, Finland
**Genotyping**
Kati Donner Institute for Molecular Medicine Finland, HiLIFE, University of Helsinki, Finland
Timo P. Sipilä Institute for Molecular Medicine Finland, HiLIFE, University of Helsinki, Finland
**Sample Collection Coordination**
Anu Loukola Helsinki Biobank / Helsinki University and Hospital District of Helsinki and Uusimaa, Helsinki

**Sample Logistics**

| | |
|---|---|
| Päivi Laiho | THL Biobank / The National Institute of Health and Welfare Helsinki, Finland |
| Tuuli Sistonen | THL Biobank / The National Institute of Health and Welfare Helsinki, Finland |
| Essi Kaiharju | THL Biobank / The National Institute of Health and Welfare Helsinki, Finland |
| Markku Laukkanen | THL Biobank / The National Institute of Health and Welfare Helsinki, Finland |
| Elina Järvensivu | THL Biobank / The National Institute of Health and Welfare Helsinki, Finland |
| Sini Lähteenmäki | THL Biobank / The National Institute of Health and Welfare Helsinki, Finland |
| Lotta Männikkö | THL Biobank / The National Institute of Health and Welfare Helsinki, Finland |
| Regis Wong | THL Biobank / The National Institute of Health and Welfare Helsinki, Finland |

**Registry Data Operations**

| | |
|---|---|
| Hannele Mattsson | THL Biobank / The National Institute of Health and Welfare Helsinki, Finland |
| Kati Kristiansson | THL Biobank / The National Institute of Health and Welfare Helsinki, Finland |
| Susanna Lemmelä | Institute for Molecular Medicine Finland, HiLIFE, University of Helsinki, Finland |
| Sami Koskelainen | THL Biobank / The National Institute of Health and Welfare Helsinki, Finland |
| Tero Hiekkalinna | THL Biobank / The National Institute of Health and Welfare Helsinki, Finland |
| Teemu Paajanen | THL Biobank / The National Institute of Health and Welfare Helsinki, Finland |

**Sequencing Informatics**

| | |
|---|---|
| Priit Palta | Institute for Molecular Medicine Finland, HiLIFE, University of Helsinki, Finland |
| Kalle Pärn | Institute for Molecular Medicine Finland, HiLIFE, University of Helsinki, Finland |
| Shuang Luo | Institute for Molecular Medicine Finland, HiLIFE, University of Helsinki, Finland |
| Vishal Sinha | Institute for Molecular Medicine Finland, HiLIFE, University of Helsinki, Finland |

**Trajectory Team**

| | |
|---|---|
| Tarja Laitinen | Pirkanmaa Hospital District, Tampere, Finland |
| Harri Siirtola | University of Tampere, Tampere, Finland |
| Javier Gracia-Tabuenca | University of Tampere, Tampere, Finland |
| Mika Helminen | University of Tampere, Tampere, Finland |
| Tiina Luukkaala | University of Tampere, Tampere, Finland |
| Iida Vähätalo | University of Tampere, Tampere, Finland |

**Data protection officer**

| | |
|---|---|
| Tero Jyrhämä | Institute for Molecular Medicine Finland, HiLIFE, University of Helsinki, Finland |

**FinBB - Finnish biobank cooperative**

Marco Hautalahti
Laura Mustaniemi
Mirkka Koivusalo
Sarah Smith
Tom Southerington

#### Biogen Biobank Team contributors

**Steering team:** Ellen Tsai, Christopher D. Whelan, Paola Bronson, David Sexton, Sally John, Heiko Runz.
**Data management team:** Eric Marshall, Mehool Patel, Saranya Duraisamy, Timothy Swan.
**Extended scientific team:** Dennis Baird, Chia-Yen Chen, Susan Eaton, Jake Gagnon, Feng Gao, Cynthia Gubbels, Yunfeng Huang, Varant Kupelian, Kejie Li, Dawei Liu, Stephanie Loomis, Helen McLaughlin, Adele Mitchell, Nilanjana Sadhu, Benjamin Sun, Ruoyu Tian.
