## Extended Data Figures for "Genetic associations of protein-coding variants in human disease"

### Extended Data Figure 1. UKB and FG study overview

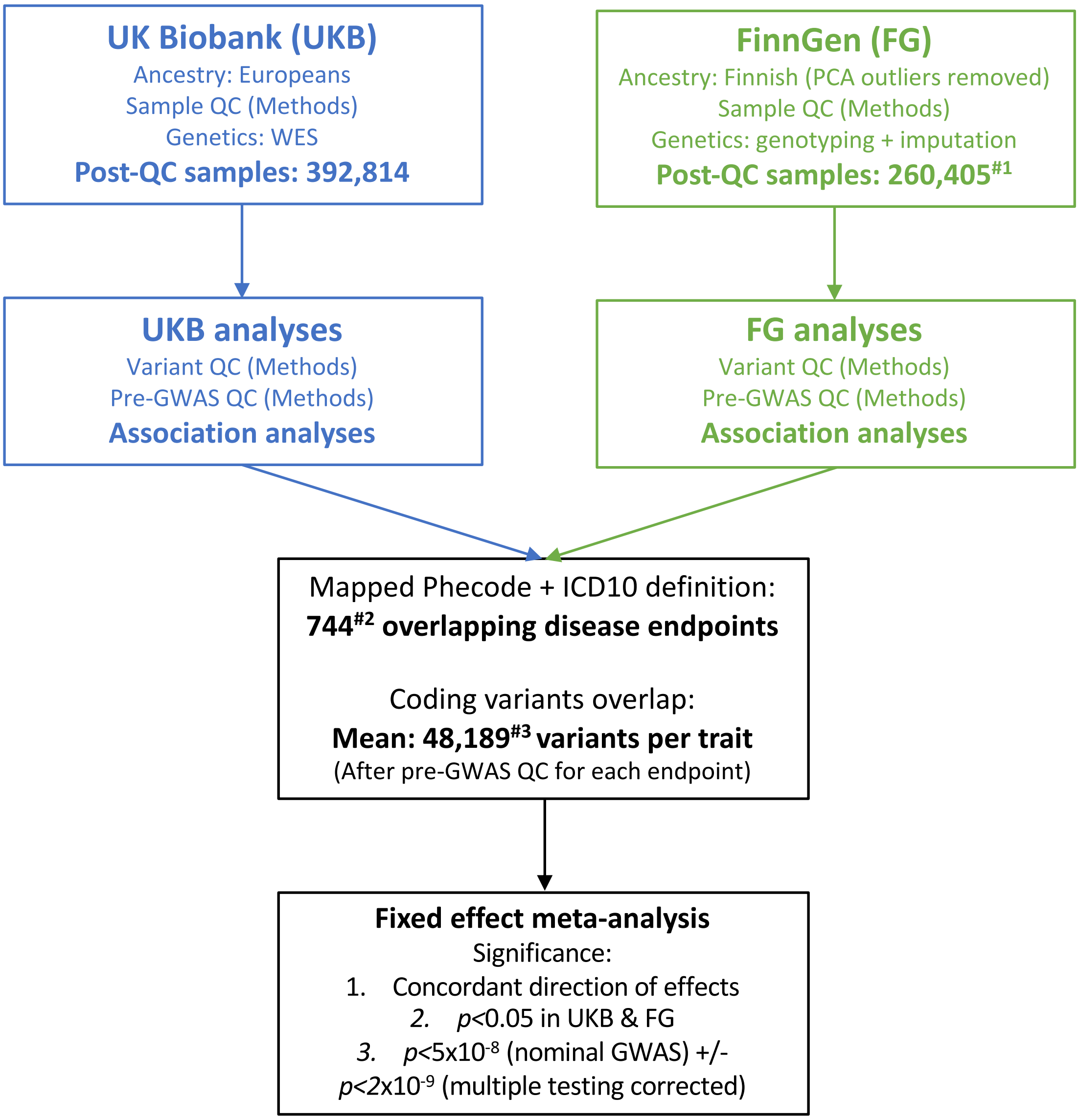


[#1]R5 of chip-based genotyping data (n=150,831), R6 of imputed data (n=260,405)
[#2]only included matching (score≥0.7) endpoints with cases ≥ 100
[#3]variants vary after filtering for MAC≥5 and MAC≥3 in cases

**Extended Data Figure 2. Case count comparison between UKB and FG across disease groups**

Diseases within each group are listed in **Supplementary Table 2**. Only cases >100 in UKB/FG are included. *R: Spearman's correlation for FG R5 (red) and R6 (blue).*

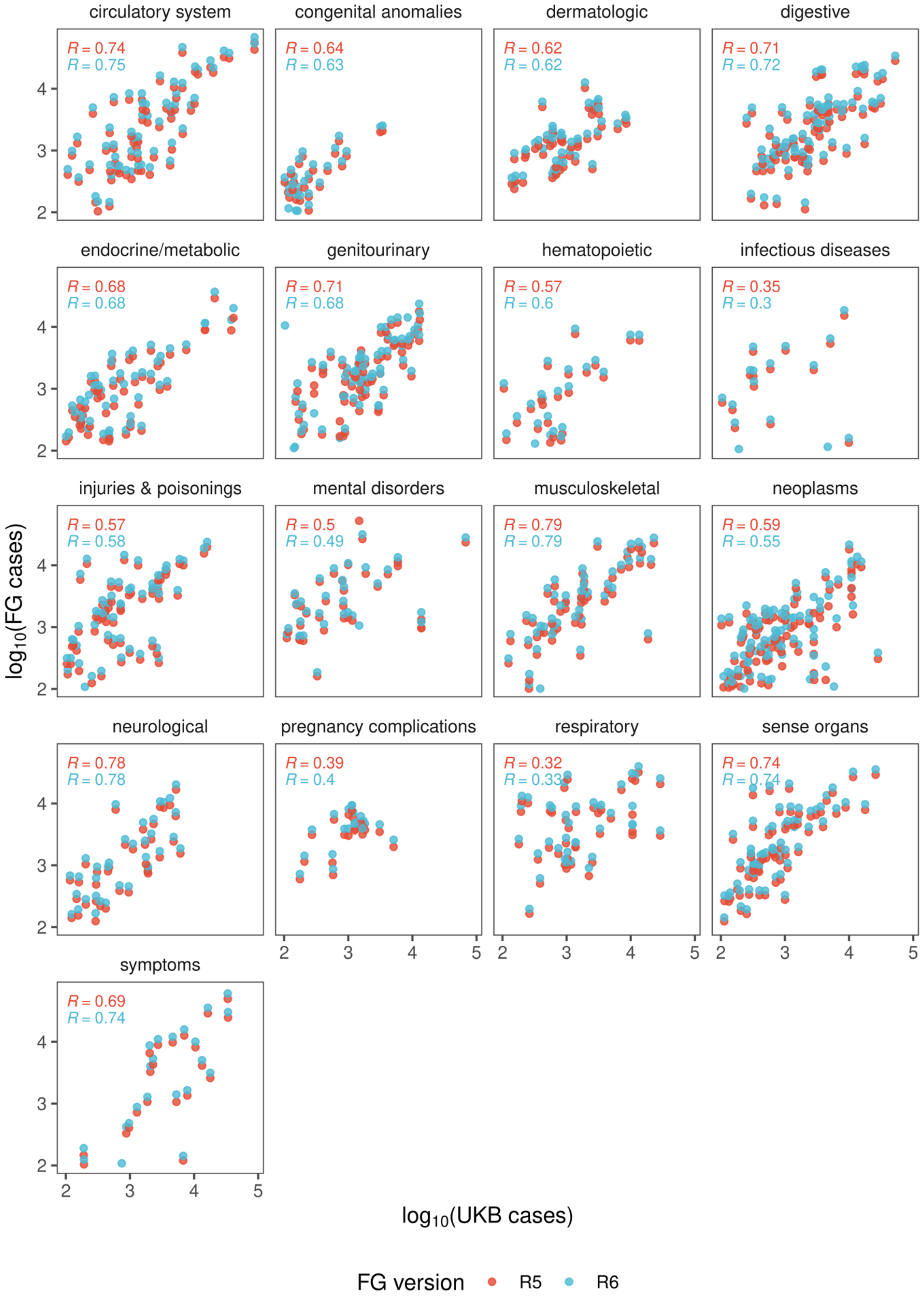

**Extended Data Figure 3. Distribution of variant annotation categories. Left**: all variants tested. **Right**: variants with at least 1 significant association ($p$<5x10$^{-8}$).
pLOF: predicted loss of function. LC: low confidence loss of function.

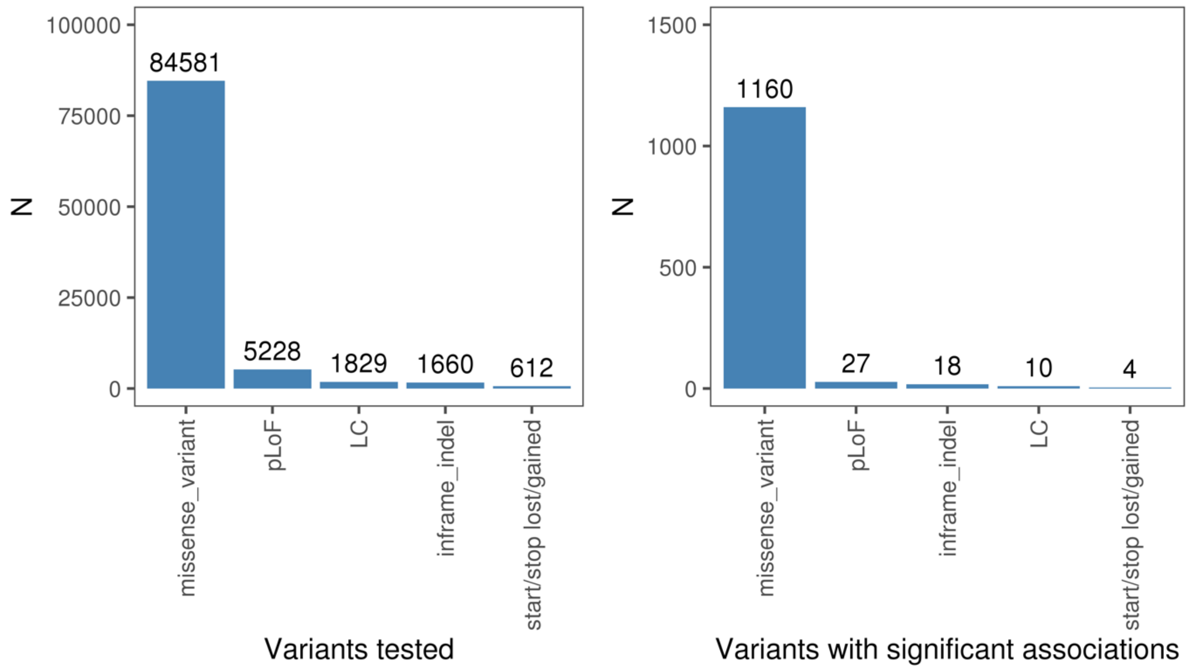

**Extended Data Figure 4. Inflation factors and FG-UKB effect size comparisons**
**(a) Distribution of inflation factors of CWAS meta-analysis. (b) Effect size comparison between UKB and FG.** Inset: zoomed in on small effect sizes. *R: Spearman's correlation*

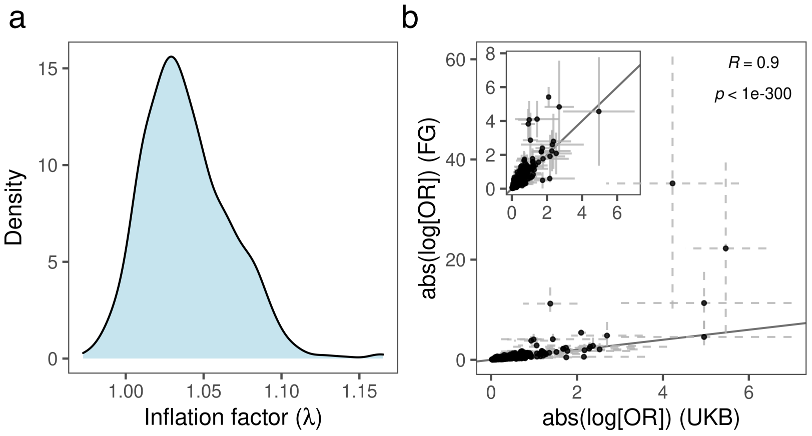

**Extended Data Figure 5. Surface plot of effects of cohort specific allele enrichment on inverse variant weighted meta-analysis z-scores (IVW uplift) across MAFs** (up to MAF 1%). Uplift is defined as the ratio of meta-analysed IVW Z-score to the Z-score of an individual study. **Left:** theoretically predicted IVW uplift. **Middle**: observed IVW uplift. **Right**: Median absolute relative error (MARE, %) between simulated and theoretical IVW uplift values**.** For each combination of MAF and allelic enrichment, we simulated 1000 datasets for two binary variables reflecting disease status for two studies. Study sample size and disease prevalence were fixed (matching values estimates from UKB and FG), genomic effects were randomly sampled from the set of positive effect sizes in UKB and FG (**Supplementary Table 3**), MAF was varied from 0.01% to 1% and allele enrichment (in the smaller study) ranged from 1 to 50.

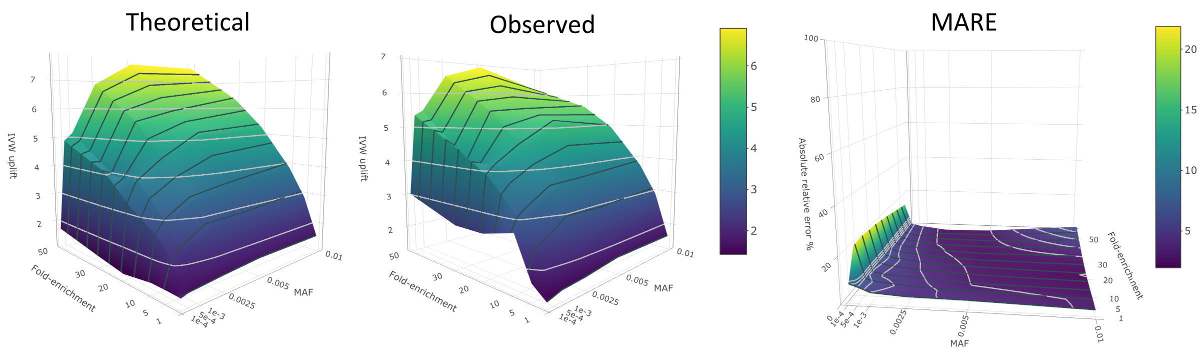

**Extended Data Figure 6. Histogram of disease and biomarker associations per region.** **(a) Number of associated trait clusters ($p$<5x10$^{-8}$) per region.** Inset shows zoomed in x-scale between 0-12 trait cluster associations per region. **(b) Number of associated biomarker groups per locus ($p$<1x10$^{-6}$).**
*MHC: Major Histocompatibility Complex.*

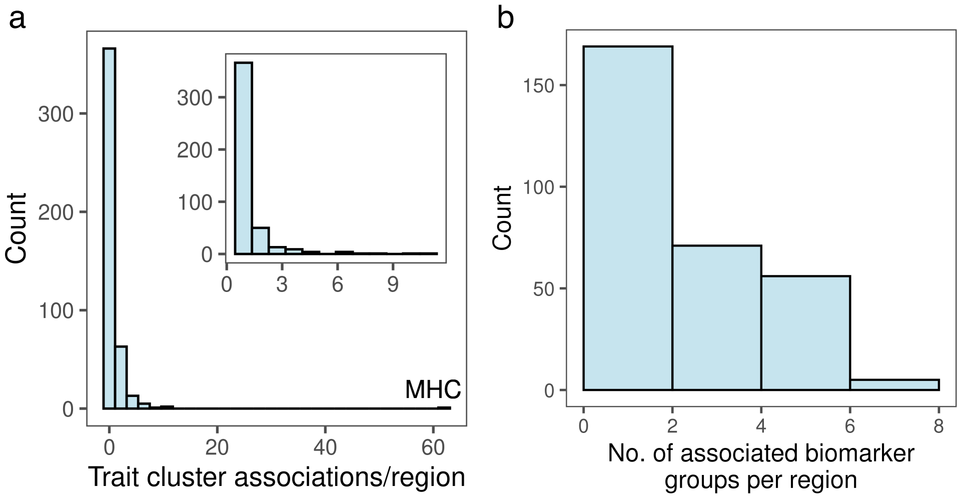
